# Decoding ruminative states from neurophysiological patterns

**DOI:** 10.1101/2024.05.15.24307414

**Authors:** Jana Welkerling, Patrick Schneeweiss, Sebastian Wolf, Tim Rohe

## Abstract

Individuals with depression often engage in iterative “rumination” about challenging situations and potential outcomes. Although the state of rumination has been associated with diverse univariate neurophysiological features, the potential to use multivariate patterns to decode it remains uncertain. In this study, we trained linear support vector machines to differentiate state rumination from distraction using patterns in the alpha, beta, and theta bands, as well as inter-channel connectivity. We used validated tasks to induce rumination or distraction for eight minutes in 24 depressed individuals in six runs over three sessions. During inductions, we recorded 64-channel EEG data and measured self-reported levels of rumination. Participants reported strongly increased rumination, and we decoded state rumination from EEG patterns with significant accuracy. However, the informative features were not consistent across participants, demonstrating that while ruminative states can indeed be decoded from EEG data, these states appear to reflect processes unique to each individual.

## 1 Introduction

When faced with an emotionally challenging situation, such as social exclusion, humans often start to think about the situation, potential solutions, and their own negative emotional reaction to it. This process of “chewing” on negative thoughts long beyond the eliciting situation is known as “rumination” and has been defined as “a mode of responding to distress that involves repetitively and passively focusing on symptoms of distress and on the possible causes and consequences of these symptoms” (Nolen-Hoeksema, Wisco, & Lyubomirsky, 2008). Typically, ruminative thoughts are accompanied by negative emotional reactions. While some situations induce rumination in most people, there is variability in individuals’ general tendency to ruminate. Thus, rumination can be conceptualized both as a transient state experienced in response to a distressing situation and as a stable personality trait.

Trait rumination is a key clinical feature of depression, as well as other psychiatric disorders such as anxiety and eating disorders (Aldao, Nolen-Hoeksema, & Schweizer, 2010). Depression, one of the most burdensome diseases worldwide (GBD 2019 Mental Disorders Collaborators, 2022), is characterized by persistent low mood, loss of interest, and low energy. Trait rumination is linked to the onset of depression, acting as a risk factor (Aldao et al., 2010; Kuyken, Watkins, Holden, & Cook, 2006; Watkins & Roberts, 2020), as well as to the duration and the severity of depression, with a stronger tendency to ruminate being associated with longer episodes (Nolen-Hoeksema, 1991; Timm et al., 2017) and an increased risk of suicide attempts (Chiang et al., 2022; Eshun, 2000).

Individuals who exhibit heightened ruminative thought patterns during emotionally challenging situations are more likely to demonstrate higher levels of trait rumination (Fang, Marchetti, Hoorelbeke, & Koster, 2019). In fact, neurocognitive models, such as the attentional scope model of rumination (Whitmer & Gotlib, 2013), have identified state rumination as a key cognitive factor that perpetuates momentary depressive symptoms (Timm et al., 2017; Watkins & Roberts, 2020) through a vicious cycle: When in a sad mood, individuals with depression narrow their attentional focus to their mood, its causes, and consequences, impeding the retrieval of alternative thoughts. By maintaining information related to the sad mood in their working memory, patients are unable to access anti-depressive strategies stored in their long-term memory, thus perpetuating their sad mood. To disrupt this cycle of repetitive negative thoughts and feelings, individuals attempt to interrupt rumination, either physiologically, for example by abusing alcohol (Devynck, Rousseau, & Romo, 2019), or psychologically, by distracting themselves from negative thoughts. Employing distraction as a cognitive coping strategy involves deliberately redirecting one’s attentional focus from the distressing thoughts towards neutral or enjoyable activities (Nolen-Hoeksema, 1991), offering transient relief from the sad mood.

In individuals with depression, rumination appears to be a highly stable recursive attractor state of brain dynamics (Rolls, 2016) accompanied by specific cognitive and negative affective mental representations. Rumination has been associated with numerous aspects of brain dynamics, such as activity in the default mode network as observed through fMRI (Zhou et al., 2020) and neural oscillations indicating functional connectivities as evident in spectral EEG features (see below). However, it is unknown whether an experimentally induced ruminative brain state, as measured by neurophysiological patterns, can be distinguished from alternative brain states such as distraction. Should this be achievable, it would enable the classification of an individual’s ruminative state based on his or her dynamic brain activity. This capability would not only contribute to neurocognitive (e.g., Whitmer & Gotlib, 2013) and clinical models of the neural dynamics of state rumination, but also offer a means for measuring ruminative states through neurophysiological data online e.g., during intervention studies.

In this study, we therefore aimed to decode state rumination from neurophysiological EEG patterns as a proof-of-principle. We chose EEG for its ability to capture brain dynamics with high temporal resolution, presenting a more practical option than fMRI, which is also often not feasible in interventional studies, such as those involving exercise. EEG studies have linked both trait and state rumination with numerous univariate neurophysiological features. Trait rumination has been shown to be positively associated with functional beta-band connectivity between the posterior cingulate cortex (PCC) and subgenual prefrontal cortex (sgPFC) (Benschop et al., 2021). Resting-state EEG measurements have demonstrated that higher levels of trait rumination are associated with asymmetrically increased alpha power over the left parietal-occipital cortex (Umemoto et al., 2021) and decreased alpha power over the right frontal cortex (Keune, Bostanov, Hautzinger, & Kotchoubey, 2011). To investigate neurophysiological correlates of state rumination, previous studies, such as Laicher et al. (2022) and Huffziger and Kuehner (2009), have experimentally induced rumination (mostly in contrast to distraction) in depressed patients to elicit increased self-reported rumination and negative affect. These studies have demonstrated that state rumination correlates with higher EEG beta power in the left temporal cortex compared to a positive and a neutral condition (Ferdek, van Rijn, & Wyczesany, 2016), and with an increased EEG alpha power over the prefrontal cortex (Putnam & McSweeney, 2008). Moreover, internal attention, which is closely linked to rumination (Magosso, Ricci, & Ursino, 2021), was found to increase EEG theta power over frontal midline regions. External attention, however, which is closely linked to distraction, was found in the same study to increase theta power while decreasing alpha power over parieto-occipital regions (Magosso et al., 2021).

Overall, the body of research examining the neurophysiological correlates of state rumination through EEG is very limited, with most studies to date having focused on trait rumination. Those studies that have investigated state rumination have been subject to methodological shortcomings, such as the absence of self-reported state rumination measurements (Ferdek et al., 2016). Moreover, prior studies examining the neurophysiological correlates of state rumination have used group-based analysis. Such an approach may not be ideal given the results of meta-analyses of brain activation patterns associated with depression, which have shown a lack of statistical significance when aggregated across individuals (Horato, Quagliato, & Nardi, 2022). Thus, it seems very likely that complex mental states such as state rumination are characterized by substantial interindividual variability in neurophysiological patterns.

Two recent studies used a *multivariate* decoding approach to decode *trait* rumination from neurophysiological patterns that suggests that trait rumination is reflected in consistent patterns across participants: Aydın and Akın (2022) were able to distinguish between high and low trait ruminators, as defined by self-report questionnaires, through spectral EEG patterns. Similarly, Kim et al. (2023) predicted *trait* rumination, as measured by a rumination questionnaire, from connectivity patterns in resting-state fMRI data (Kim et al., 2023). However, while both of these studies successfully trained their *trait rumination* decoding models on data aggregated *across* participants, they did not consider individual variability in the mapping between rumination and neurophysiological patterns. Additionally, it remains unknown whether *state* rumination is associated with neurophysiological patterns unique to each individual and how temporal dynamics of state rumination behave.

In a proof-of-concept study, we therefore addressed the following research questions: 1) Do inductions of rumination or distraction lead to differential brain states that can be decoded from neurophysiological patterns)? 2) Are the neurophysiological patterns encoding rumination consistent across individuals, or are they unique to each individual? 3) Are decoded and self-reported levels of rumination correlated across individuals? 4) Which features of the neurophysiological patterns are most informative for decoding state rumination? Answers to these question will not only provide fundamental insights which neurophysiological states of the brain correlate with ruminative states, but they will also provide a measurement tool to quantify rumination from neurophysiological data, for example to assess intervention effects on rumination (Welkerling et al., 2024).

To answer our research questions, we experimentally induced state rumination versus state distraction and used multivariate pattern analysis to decode the two states from neurophysiological patterns (Fig. 1). During the inductions, we collected EEG data and measured self-reported rumination and affect before, during, and after the inductions. To classify rumination and distraction, we trained support vector machines (SVM) to decode both states from spectral EEG features in the alpha, beta, and theta bands, as well as functional connectivity, which have been linked to rumination in previous research. As alpha and beta connectivity have been linked to rumination, we decided to calculate whole-spectrum connectivity to ensure that no potentially relevant information, such as from other frequency bands, is overlooked. (Benschop et al., 2021; Ferdek et al., 2016; Keune et al., 2011; Magosso et al., 2021; Putnam & McSweeney, 2008; Umemoto et al., 2021).

**Figure 1.**
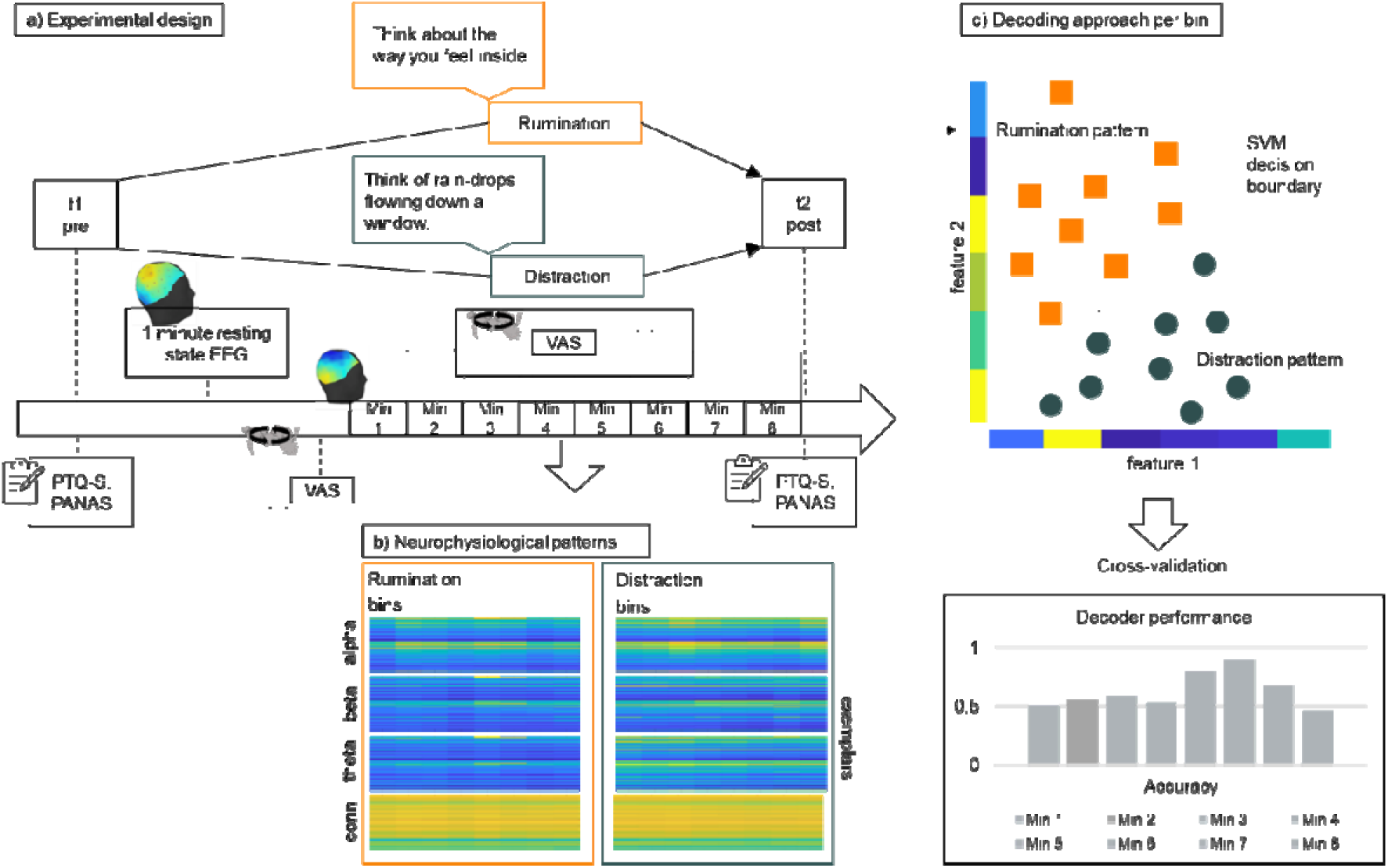
Experimental design and decoding approach. a) The experimental design measured self-reported rumination and affect (i.e., Perseverative Thinking Questionnaire (PTQ-S),Positive and Negative Affect Schedule (PANAS); for further details please refer to the Methods section), as well as one-minute resting state EEG data, before visual items induced rumination or distraction. During the eight-minute inductions, participants reported their ruminative and affective state on a seven-point visual analogue scale (VAS) every minute. b) To decode rumination versus distraction from neurophysiological patterns, we computed feature vectors comprising alpha (60 features), beta (60 features), and theta (60 features) power, as well as connectivity between electrodes (1770) from eight one-minute bins of 64-channel EEG data. c) Next, we trained linear SVM decoders to distinguish between induced rumination and distraction from neurophysiological feature vectors (feature values are shown on the x and y axis and exemplars are the dots lying in x/y space) using cross-validation. To evaluate decoder performance in each one-minute bin, we computed the proportion of correctly predicted state labels.

## 2 Methods

### 2.1 Participants

Twenty-four moderately or severely depressed individuals (17 female, *M_age_* = 24.6 years) were included in the study after providing written informed consent. A pilot study with *n* = 6 participants was conducted to calculate statistical power and sample size for the main study. For a proof-of-concept study such as ours, the primary goal was to show that decoding accuracy is significantly larger than a chance level of 50%. Thus, we converted decoding accuracy from the pilot sample into Cohen’s d to compute the required sample size to statistically safeguard above-chance decoding accuracy. In the pilot study the mean decoding accuracy across all participants differed with a medium effect size (*d* = 0.60) from chance-level. Based on this effect, power analysis using the open-source programme G*Power (3.1) resulted in a minimum sample size of 24 people (two-tailed *t*-test for dependent samples, _α_-level of .05, power of 80%, not taking into account possible drop-outs). In addition to the six data sets that had already been collected, data from an additional 18 participants were also gathered. The pilot study additionally involved a positive mood induction and fNIRS measurements. Because there were no other changes in the experimental design or setup between the pilot and main studies, we included data from these six individuals in the main sample. Sociodemographic and clinical information of the participants can be found in Tab. S10. Sensitivity analyses including only the main sample are described in the supplementary results (Fig. S4).

Inclusion criteria were a diagnosis of moderate or severe depression according to the Structured Clinical Interview for DSM-5 Disorders, Clinician Version (SCID-V; Beesdo-Baum, Zaudig, & Wittchen, 2019); F32.1-F32.2, F33.1-F33.2 DSM-V), a Beck Depression Inventory II (BDI-II; Beck, Steer, & Brown, 1996) score greater than 19, age between 18 and 40 years, fluency in German, no current psychopharmacological treatment, and no current psychological treatment. To ensure that rumination and distraction inductions reliably altered the mental states in each participant, we only included participants who showed an increase in rumination after a rumination induction pretest (PTQ-S_post-pre_ > 0) and a decrease in rumination after the distraction induction pretest (i.e., PTQ-S_post-pre_ < 0). Exclusion criteria were acute suicidality, schizophrenia or other psychotic disorders (F23, F20.81, F20.9 DSM-V), acute substance misuse excluding tobacco (F10, F11, F12, F13, F14, F15, F16, F18, F19 DSM-V), acute eating disorders (F50.01, F50.02, F50.2, F50.81, F50.82 DSM-V) acute or chronic diseases or conditions influencing brain metabolism, such as diabetes mellitus (E10-E14 ICD-10), kidney insufficiency of stage 3 according to the Kidney Disease Outcome Quality Initiative Guidelines (National Kidney Foundation, 2002), non-adjusted hypertension (I10 ICD-10), and moderate to severe craniocerebral trauma (Glasgow Coma Scale; Teasdale & Jennett, 1974; GSC 3-12) or craniocerebral trauma second or third degree with loss of consciousness for more than 30 minutes. Additionally, we excluded participants who engaged in regular physical exercise, as this was an exclusion criterion pertinent to the second part of the study, details of which will be reported in a separate publication (Welkerling et al., 2024); for more information, refer to our pre-registration document available online at <u>osf.io/qf2ha</u>. Participants were recruited through local hospital departments and ambulatory psychotherapeutic practices. Participants were compensated financially and provided with additional mental health treatment information. The study was approved by the local Ethics Committee for Medical Research of the University of Tübingen (133/2020BO2) and was conducted in accordance to the tenets of the Declaration of Helsinki of 2010.

### 2.2 Procedure

Individuals interested in taking part in our study were first screened for eligibility in a telephone call based on the pre-defined inclusion and exclusion criteria. Those found eligible were then invited to an in-person appointment. After providing informed written consent, they underwent a clinical interview (SCID-V) conducted by a psychologist trained in clinical diagnostics. They also completed questionnaires to measure the severity of their depression (BDI-II) and their general tendency towards rumination (PTQ). Additionally, this initial appointment included pretests for the rumination and distraction induction protocols to determine if these procedures led to a sufficient change in individuals’ levels of rumination, as required for inclusion in the study. At a subsequent appointment, participants were medically examined by sports physicians for eligibility to engage in the physical exercises involved in the second part of the study, which will be reported in a separate publication (Welkerling et al., 2024). In the second week of the study, participants attended three sessions during which they underwent both rumination and distraction inductions consecutively to generate training data for the decoder. The order of these inductions was randomized within each session but counterbalanced across participants.

### 2.3 Experimental tasks

Before each induction, a baseline resting-state measurement was taken, with participants closing their eyes for 60 seconds while EEG data were collected. For the rumination induction protocol, participants were exposed to 16 different items, which were adapted from Huffziger and Kuehner (2009) and designed to direct participants’ attention towards their own symptoms and emotions (e.g., “Think about the way you feel inside.”). Each item appeared on the screen for seven seconds, and participants were instructed to reflect on it. Following this, a blank screen was displayed for 30 seconds to avoid reading artifacts, and participants were asked to continue focusing on the item. After every two items, participants assessed their level of rumination and their affective state using two consecutive visual analogue scales (VAS; see supplemental methods for VAS). The VAS appeared on the screen and participants rated their current levels of rumination and affect by pressing the corresponding button on the keyboard. Before the induction protocols started, participants were familiarized with the definition of rumination. We explicitly instructed participants that rumination does not refer to general thinking, but rather means brooding about their own emotions and symptoms.

For the distraction induction protocol, participants were exposed to 64 items from four different tasks, all designed to shift their attentional focus away from their own symptoms and distress. In the first task, adapted from Huffziger and Kuehner (2009), 16 different prompts (randomly selected from the original set of 28 items) were presented, aiming to elicit neutral mental images (e.g., “Think of raindrops flowing down a window”). The second task was a knowledge-based exercise, requiring participants to evaluate the correctness of 16 randomly selected statements from the Distraction Questionnaire by Ehring, Fuchs, and Kläsener (2009) (e.g., “Spain is not a neighboring country of Germany!”), with the response options “true” and “false”. The third task, also adapted from Ehring et al. (2009), was a word-generation activity requiring participants to think of a new word beginning with each letter from given five-letter words (e.g., from “ocean”: oven, concentration, elephant, apple, night). The fourth task, developed by the authors of this paper, aimed to redirect participants’ attention towards self-related topics unrelated to symptoms, similar to the prompts of Huffziger and Kuehner (2009) (e.g., “Think about your next weekly shopping”). In each trial, an item from one of the four tasks was presented for two to five seconds, with their order randomized within and across participants. After each item, a blank screen appeared for 7.5 seconds. Participants were instructed to maintain their focus on the previously presented item. After every eight items, participants reported their level of rumination and affective state using the VAS. Before and after each induction, levels of rumination and affect were assessed with the PTQ-S and the PANAS, respectively. Further details concerning the PANAS and VAS affect can be found in the supplementary methods. To accommodate potential shifts in attentional focus away from the screen, a beep was used to signal the onset of the next item or the VAS (Fig. 1).

The experiments were conducted in a laboratory room using Psychtoolbox 3.0.16 (Brainard, 1997) running under MATLAB 2020b (The MathWorks Inc., 2020) on a laptop connected to an LCD screen (Acer B246HYL; 60.5 cm diagonal display size). Acoustic stimuli were delivered at approximately 77 dB SPL through speakers (Amazon basics V216Custom1) positioned to the right and left of the screen. Participant responses were collected with a keyboard connected to the laptop via USB. Participants were seated approximately 75 cm away from the screen.

### 2.4 Self-report measurements

#### 2.4.1 Perserverative Thinking Questionnaire (PTQ)

The German version of the PTQ comprises 15 items to assess the core processes of repetitive negative thinking: repetitive, intrusive, difficult to disengage from, unproductive and capturing mental capacity. Nine items load on the first three processes and three on each of the last two. Furthermore, a total sum score for all 15 items can be computed and assesses the general tendency to ruminate. In the PTQ-S, the state version, instructions change to how participants thought about negative events and problems in the last moments. Higher values correspond to higher levels of rumination (Ehring et al., 2011). Compared to the widely used Response Styles Questionnaire (RSQ) by Nolen-Hoeksema and Morrow (1991), the PTQ focuses on key processes of rumination without specifying the content of those thoughts. Since we were interested in the underlying processes of rumination compared to distraction, we measured rumination with the PTQ. Internal consistency for the total sum score amounted to α = .95 in a clinical sample and construct validity to the RSQ amounted to *r* = .72 (Ehring et al., 2011). Of a total of 288 timepoints seven were missing (2.43%). Missing values were handled by averaging the total sum score of the PTQ-S per induction and timepoint over the three sessions.

#### 2.4.2 Visual Analogue Scales (VAS)

Adapted to the self-assessment manikins from Bradley and Lang (1994) the authors designed manikins assessing rumination and affect. The VAS range from 1 to 7. For rumination one corresponds to *“no rumination”* and seven to *“a lot”* (see supplemental methods). 54 from 1296 datapoints of self-reported VAS-rumination were missing (4.17%). Missings were handled by averaging the VAS responses per induction and timepoint over the three sessions.

### 2.5 EEG measurements

EEG data were recorded using a 64-electrode actiCHamp Plus system (Brain Products GmbH, Gilching, Germany) with a sampling rate of 1000 Hz. Fifty-eight of these electrodes were positioned according to the international 10-20 system (Jasper, 1958). Five of the 64 electrodes (i.e., O1, O2, TP9, TP10, T8) deviated from the 10-20 system and were placed around the eyes and mouth to monitor eye movement and facial motion (adjacent to both the right and left eyes; below and above the left eye; on the risorius muscle). FCz served as the reference and AFz as the ground electrode. Additionally, to control for muscle artifacts from neck and shoulder movements, four pairs of bipolar electrodes were applied: two pairs on either side of the trapezius muscle, and two pairs on either side of the neck targeting the sternocleidomastoid muscles. To monitor head movements, an accelerometer was attached to the electrode cap to the left of the Cz electrode. In the case of the six pilot study participants, 59 electrodes were placed according to the 10-20 system. However, for the pilot participants, a separate reference electrode channel was used, which was part of the electrode setup for the main sample: for them, the Iz electrode was placed in FCz, which then served as the reference electrode in the main sample.

#### 2.5.1 EEG preprocessing

The pre-processing of EEG data was carried out using Brainstorm software (Tadel, Baillet, Mosher, Pantazis, & Leahy, 2011) (version 14 October 2022) in six steps. First, to remove muscle-and head-motion artifacts, the raw EEG data were corrected for muscle and movement artifacts using sliding-window regressions (van der Meer et al., 2016) implemented in EEGLab (Delorme & Makeig, 2004) (O1, O2, TP9, TP10, T8, bipolar electrodes from the trapezius and sternocleidomastoid muscles, as well as the accelerometer). Second, the EEG data were bandpass filtered with a frequency range of 0.25 to 45 Hz. Third, eye blinks were automatically detected by Brainstorm’s standard algorithms using data from the electrode above the left eye. Signal-space projectors (SSPs) were subsequently created from 400 ms segments, band-pass filtered between 1.5 and 15 Hz and centered on the detected blinks. The first spatial component of the SSPs was then used to correct for blink artifacts in the continuous EEG data. Fourth, all data were visually inspected to identify and exclude any segments showing artifacts from blinks, saccades, motion, electrode drifts, or jumps. Fifth, whole channels were marked as bad if visual inspections revealed suspicious time courses or spectral power distributions resulting from artifacts. Bad channels were interpolated as the distance-weighted average of neighboring channels. Lastly, data were re-referenced to the PO7 and PO8 electrodes (PO9 and PO10 for the pilot sample) and resampled to a frequency of 250 Hz.

To ensure that EEG data used for training the decoder were not contaminated by reading artifacts, analysis was restricted to data from the blank-screen sequences. Because the shortest blank-screen sequence lasted for 7.5 seconds, all blank-screen sequences were binned into 7.5-second epochs for consistency. Because the electrode set-up differed slightly between the pilot (*n* = 6) and main (*n* = 18) samples, channels that were missing (AFz in the pilot sample and PO9 and PO10 in the main sample) were interpolated similarly to previously marked bad channels, resulting in 60 channels for all participants. To compute feature vectors for the decoding algorithm, 7.5-second epochs were transformed using a Fast Fourier transformation. Feature vectors included the power of the alpha (i.e., averaged over 8 to 13 Hz), beta (i.e., averaged over 13 to 30 Hz), and theta (i.e., averaged over 4 to 7 Hz) bands across the 60 channels and averaged within each 7.5s epoch. Additionally, connectivity features comprised the pairwise connectivity between the 60 electrodes computed as pairwise correlation of time courses within the 7.5-second epochs (i.e., 1770 correlations per epoch: [60*60 channels – 60 values from the diagonal] / 2). Each feature vector was *z*-standardized (*zscore* function Matlab) to normalize scaling differences between feature vectors (Pereira, Mitchell, & Botvinick, 2009). Overall, the feature vector per 7.5-second epoch comprised 1950 features including 60 *z*-standardized alpha power values, 60 *z*-standardized beta power values, 60 *z*-standardized theta power values, and 1770 *z*-standardized connectivity values (Fig. 1). Shorter epochs of 3.75 seconds length did not change our results, we therefore chose 7.5 s to align the definition of features to the duration of the blank screen sequences from the distraction induction.

#### 2.5.2 Decoding procedure

To decode rumination versus distraction, we trained linear support-vector machine (SVM) classification models using LIBSVM (Chang & Lin, 2011). For training and prediction, we employed a leave-one-session-out cross-validation (CV) procedure: The SVMs learned the mapping between feature vectors and the two classes (rumination vs. distraction) using data from two of the three induction sessions. Subsequently, these models were applied to predict the class labels of the independent feature vectors from the remaining session. This CV procedure, recommended by best practice guidelines (Scheinost et al., 2019), ensured that training and testing of the decoders was performed on independent data sets with balanced classes within each cross-validation fold. Because we implemented a within-participant experimental design, the data from the induced experimental conditions (i.e., rumination vs. distraction) were independent in all three sessions at the level of individual participants. The SVMs’ nu parameter (i.e., from [0.0001,0.1:0.025:0.7]) was optimized through nested cross-validations. To explore the time course of rumination across the eight one-minute bins, the CV procedure was repeated for each bin. Importantly, the data of each bin were divided into eight 7.5-second epochs (see above) that were treated as data segments for decoding, resulting in an effective bin size of one minute. In other words, in each cross-validation fold for a bin, the decoder was trained on 32 data points (= 2 sessions x 2 inductions x 8 epochs). We chose eight one-minute bins, each consisting of eight 7.5-second epochs to align the decoding approach with the VAS reports that were requested every minute during inductions. This approach facilitated an exploratory comparison between the SVM decoding performance and the VAS reports, analyzing decoding probability estimates against the rumination scales. The decoder predicted class labels (i.e., rumination vs. distraction) and corresponding probability estimates for each class. The decoded probability estimates represented the probability *p* that a data segment belongs to a rumination state (if *p* > 0.5) or to a distraction state (if *p* < 0.5) as defined in the induction protocols for decoder training (i.e., the probability for the classes of rumination and distraction add up to one). Additionally, to establish a baseline decoding accuracy before the inductions, we applied the same CV procedure to the one-minute resting-state data collected before each induction, epoched similarly to the induction data. The chance level for the decoder performance was computed by repeating the CV procedure on randomized class labels within each bin (i.e., 1000 random allocations of the labels rumination versus distraction). Out of the 10368 data segments obtained from all participants and sessions, 447 (4.31%) were missing or excluded from the decoder training due to technical reasons. The decoder for minute 0 before the inductions served only to establish baseline performance; hence, all inferential analyses concerning the performance of the decoder excluded this one-minute baseline (Fig. 1).

### 2.6 Statistical analyses

#### 2.6.1 PTQ-S and VAS

To assess whether the inductions increased or decreased state rumination as intended, we averaged the total score of the PTQ-S across sessions for each induction type, timepoint (pre vs. post induction), and participant. Subsequently, we computed a linear mixed model with the averaged total score of the PTQ-S as the outcome variable. Fixed effects included induction (rumination vs. distraction), timepoint (pre vs. post), and their interaction. Random intercepts per participant were implemented. The random slopes per participant did not converge because the observed data were insufficient compared to what should have been estimated. Post-hoc two-sided pairwise *t*-tests based on estimated marginal means were used to assess differences in self-reported rumination levels for every type of induction. Additionally, estimated marginal means of the total scores were compared at every timepoint (pre vs. post) between both inductions. The significance levels for the post-hoc tests were adjusted using the Bonferroni correction to account for four comparisons (Fig. 2a).

**Figure 2.**
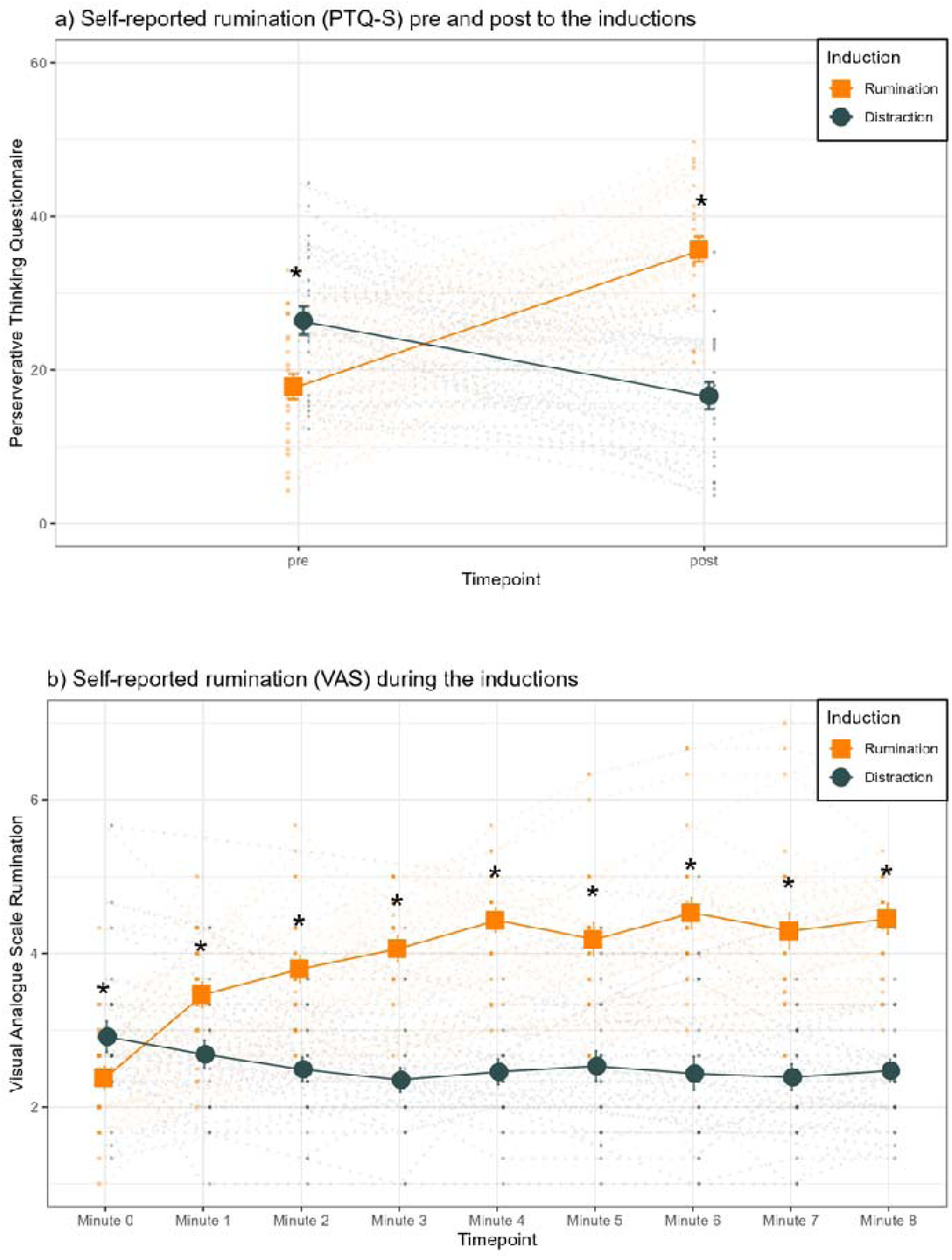
Self-reported rumination from the questionnaire and VAS scales for the rumination and distraction conditions. a) The total score of the PTQ-S for pre and post timepoints for each induction (across-participant mean +/− SEM and individual data in the background). b) Time course of VAS self-reported rumination (across-participant mean +/− SEM and individual-level data in the background). Asterisks indicate significant (*p* < .05) differences from post-hoc pairwise *t*-tests between rumination and distraction. VAS were recorded after the resting-state baseline measurement before the inductions (minute 0) and at minutes 1 to 8 during inductions (see supplemental results Tab. S1).

VAS responses were averaged across sessions for each induction type, timepoint (minute 0 to minute 8 during induction), and participant. A linear mixed model was then calculated with these averaged responses as the dependent variable. The model included fixed effects for the type of induction (rumination vs. distraction), timepoint (minute 0 to 8 during induction), and their interaction as well as random intercepts for participants. The random slopes per participant did not converge because the observed data were insufficient compared to what should have been estimated. To compare the two conditions at each timepoint, post-hoc two-sided pairwise *t*-tests based on estimated marginal means were conducted and subsequently Bonferroni corrected for nine tests (Fig. 2b). Detailed results can be found in supplementary Tab. S1 and in figure 2. Additionally, to verify whether the inductions reliably induced rumination and distraction, the averaged estimated marginal means of VAS responses over sessions for both conditions were calculated for each timepoint. Each timepoint beginning from minute 1 was compared to baseline rumination at minute 0 using a paired two-sided *t*-test (Tab. S2). These tests were also Bonferroni corrected, this time for 16 comparisons.

#### 2.6.2 Decoding analysis

To evaluate the accuracy of the person-specific decoders, the predicted class labels (rumination vs. distraction) from the independent left-out session were compared with the actual label of the data point. Decoding accuracy for each participant was quantified as a percentage of correct predictions for each bin, across all CVs and both induction types. To establish a baseline decoding accuracy for each participant in every bin, a chance level was computed by repeating the CV procedure on randomized class labels (i.e., 1000 randomized allocations of the labels to the data). Subsequently, the mean of a participant’s randomization distribution was subtracted from the original decoding accuracy for that participant to obtain chance-level-corrected decoding accuracies. For a global test of decoding accuracy, chance-level-corrected accuracies were averaged across the eight bins from the inductions for each participant, excluding the baseline measurement from the one-minute resting bin. A one-sided one-sample *t*-test was conducted to test if these chance-corrected decoding accuracies across bins and participants were significantly greater than zero. This approach allows for the assessment of whether the decoders could distinguish between rumination and distraction states beyond what would be expected by chance alone. 95%-Confidence Intervals (CI) and Bayes Factor Analysis *bf10* were calculated for the primary analysis, quantifying evidence for a difference to chance level (H□) relative to the null hypothesis of no difference (H□) using the BayesFactor toolbox (Krekelberg, 2024).

To analyze the progression of decoding accuracies throughout the eight minutes of induction, we fitted a linear mixed model with decoding accuracy as dependent variable. Bins were defined as fixed effects and random intercepts for participants were included. The random slopes per participant did not converge because the observed data were insufficient compared to what should have been estimated. To correct for multiple comparisons across bins, we derived *p*-values for the averaged accuracies across participants for each bin using randomization tests that applied the max-stat approach from Blair and Karniski (1993). In this procedure, the maximum decoding accuracies across the nine bins for each of the 1000 permutations were taken as the max-stat randomization distribution. For each bin, multiple-comparison-corrected *p*-values were then calculated as the proportion of the sum of values from the max-stat distribution that were higher than or equal to the original decoding accuracy of that bin averaged across participants (Fig. 3). Additionally, pairwise two-sided *t*-tests with the estimated marginal means of the accuracies compared accuracies of bin 0 to all other timepoints. These tests were Bonferroni corrected for eight tests.

**Figure 3.**
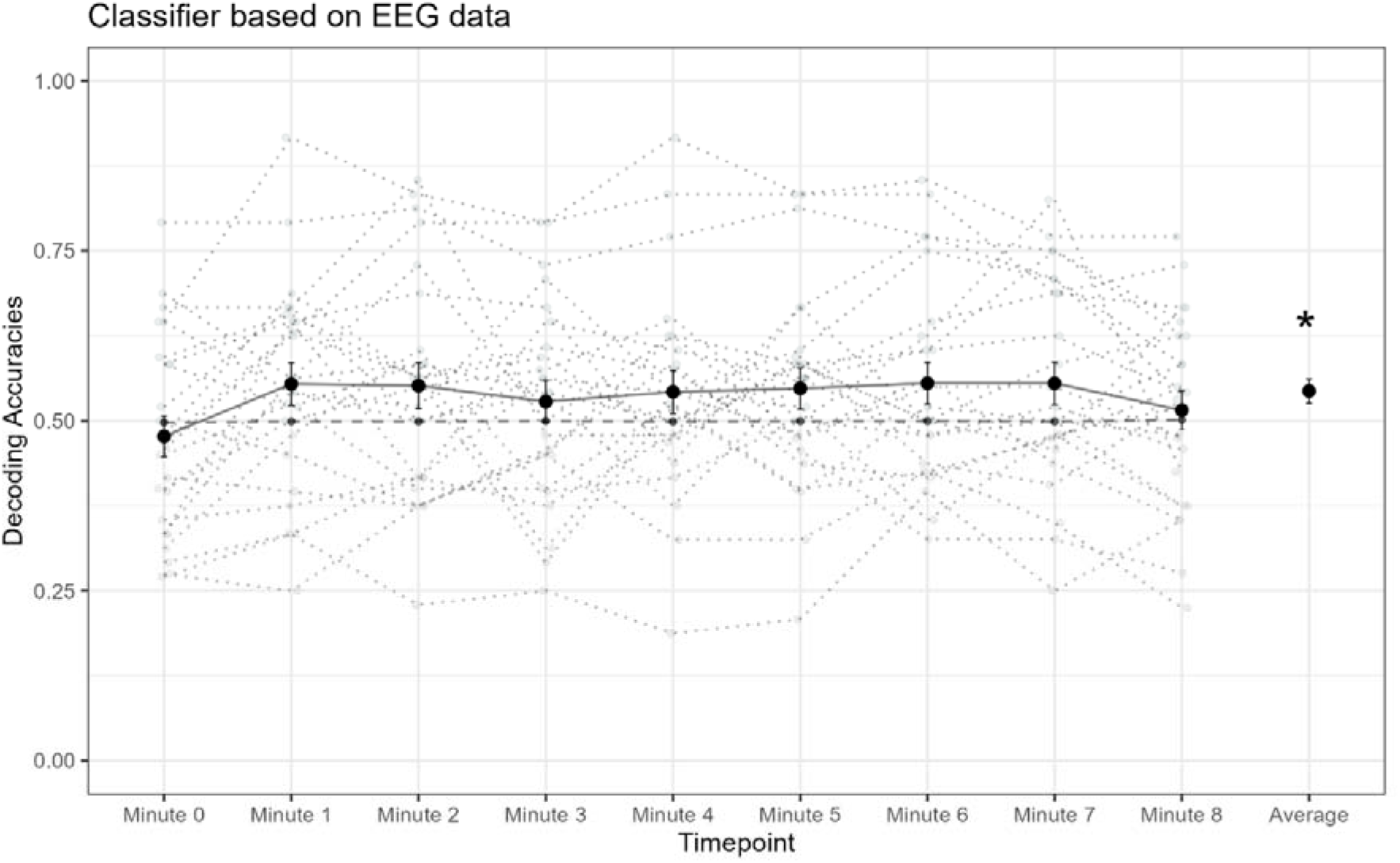
Decoding accuracies of the decoder classifying rumination versus distraction from EEG patterns across the eight-minute induction phase. Decoding accuracies (across-participant mean +/− SEM; individual data dotted in gray) for every one-minute bin and averaged over bins. Chance-level decoding accuracies (black dashed line) were calculated from randomized data (i.e., 1000 random permutations of rumination vs. distraction labels). Asterisks indicate decoding accuracies significantly larger than chance level (*p* < .05, average accuracy tested against chance-level).

For a global test of the performance of the decoder applied to each participant, we averaged the decoding accuracies across all bins for every participant and assessed whether the resulting mean decoding accuracies were significantly different from zero. This was determined using a randomization test in which the *p*-value was calculated based on the proportion of decoding accuracies from 1000 random permutations that were equal to or greater than each participant’s actual decoding accuracy (Tab. S4). All of these analyses were conducted using custom-written code in MATLAB (2020b).

Additionally, to investigate whether rumination-related EEG patterns were consistent across participants, we trained a ‘group decoder’ using combined data segments from all participants. This decoder was subjected to the same cross-validation process used for evaluating individual-level participant data. In conducting a global assessment of the accuracy of this group decoder, we averaged the decoding accuracies across the eight applicable data segments (i.e., omitting the one-minute resting period used as a temporal baseline) per participant. A one-sided one-sample *t*-test was then performed to determine if these accuracies were significantly greater than 50%, suggesting a performance above chance level.

All linear mixed models conducted for PTQ-S, VAS and decoders’ accuracies were fitted with the lmer function from the package “lme4” as well as the anova function in R (R Core Team, 2017) (version 4.4.0). Model assumptions, including linearity, normal distribution, homoscedasticity, and absence of outliers, were checked visually. Additionally for outlier analysis Cook’s Distance was calculated and if values were greater than one, sensitivity analyses were conducted as described in the following. If assumptions were violated sensitivity analyses were calculated without influential cases and if results did not differ results of the complete sample were reported. Effect sizes for the linear models were estimated using partial eta squared (Cohen, 1973) (F_to_eta2 function in R) and Cohen’s d was calculated using the R function t_to_d for post-hoc tests. Both functions belong to Rs package “effectsize”.

#### 2.6.3 Post-hoc and exploratory analysis

In our post-hoc analysis exploring the convergence or divergence of self-reported rumination and EEG-decoded rumination, we correlated the self-reported rumination levels from VAS scales and PTQ-S questionnaires with the decoders’ probability estimates of rumination versus distraction. Individual VAS values for rumination, as well as the decoders’ probability estimates, were averaged across sessions for each bin per induction type. We then performed linear correlations between the 18 VAS values (resulting from two inductions across nine timepoints) and the corresponding probability estimates. Fisher *z*-transformed individual correlations were tested against zero at the group level using one-sided one-sample *t*-tests. The analysis was complemented with the Bayes Factor *bf10*. Additionally, individual PTQ-S rumination change scores and probability estimates were averaged for each induction type and assessed through linear correlation (Fig. 4).

**Figure 4.**
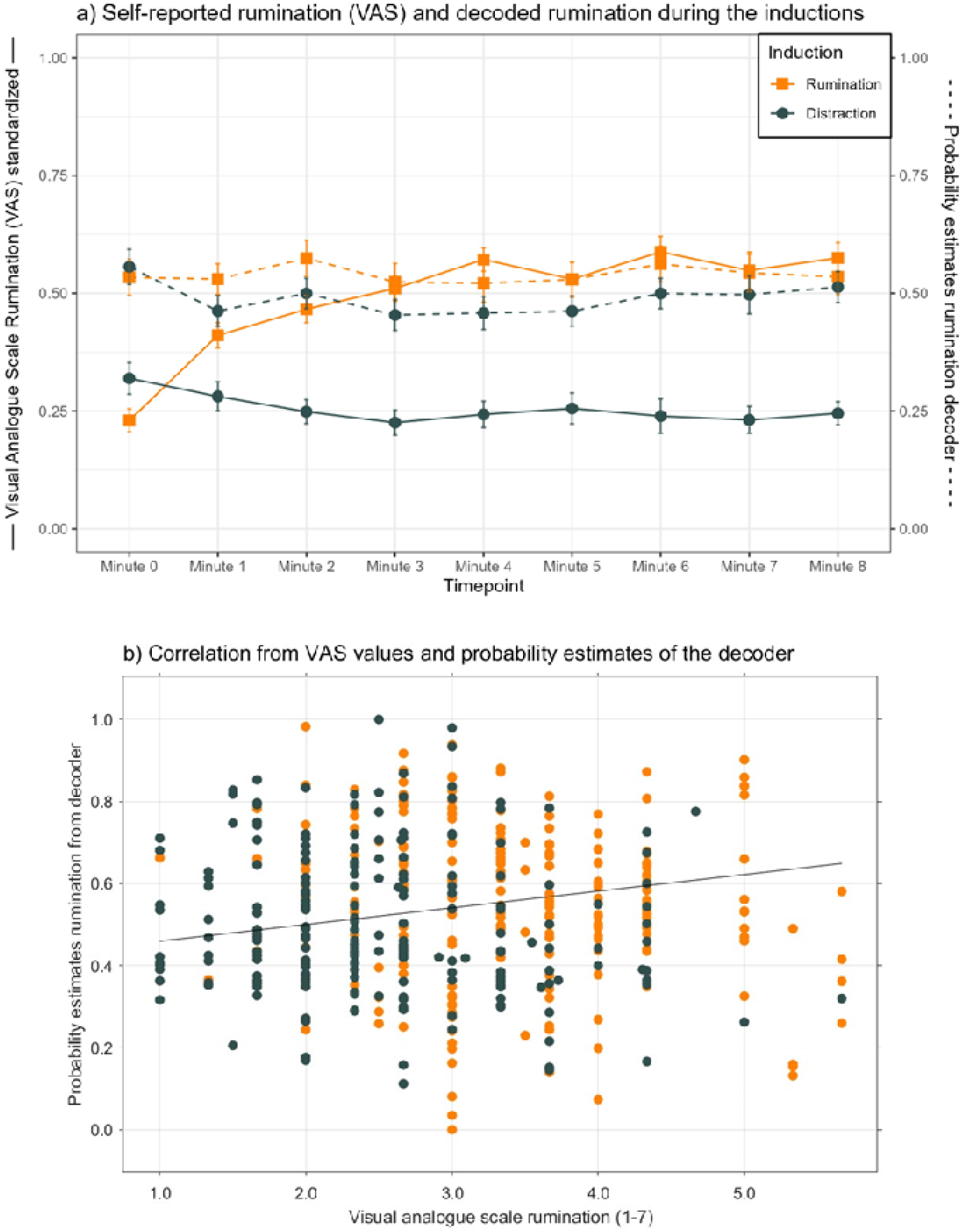
Neurobehavioral link between decoded rumination and reported rumination. a) Self-reported rumination measured with the VAS (solid lines) and decoders’ probability estimates for rumination (dashed lines) across timepoints of both induction phases. VAS values, which range from 1 to 7, were standardized between 0 and 1 (divided through six). b) Self-reported VAS rumination and decoders’ probability estimates over inductions and timepoints from all individuals. The averaged correlation between self-reported VAS rumination and decoded rumination (*r* = .24) is shown as an average regression slope.

Adopting the “virtual lesion approach” developed by Kohoutová et al. (2020) and applied by Kim et al. (2023), we assessed the contribution of different feature sets (i.e., alpha, beta, theta band power, and connectivity) to decoder performance for those participants with significant decoders (*n* = 10): First, we trained decoders excluding one feature set at a time to observe the impact on decoding accuracy compared to using the full feature sets. Subsequently, we trained a decoder on only a single feature set to assess the feature set’s decoding performance in isolation. A change in decoding performance related to the omission or isolated use of a feature set indicates the amount of rumination-related information represented by that feature set (Tab. 1). For the “virtual lesion” approach, decoders were trained as described above and baseline resting-state data were not included in the performance statistic. Comparisons between decoder performance with one feature set omitted and the full feature set decoders were made using paired two-sided *t*-tests, adjusted for multiple comparisons with Bonferroni correction for four tests. Similarly, the performance of decoders trained on single feature sets was compared to a chance level of 50% using paired one-sided *t*-tests, Bonferroni corrected for four tests. Bayes Factor Analysis (*bf10*) was additionally conducted for all *t*-tests.

**Table 1.** Results of a virtual lesion approach (Kohoutová et al., 2020) to attribute decoding performance to feature sets from the alpha, beta, and theta bands, as well as connectivity.

| Feature Set | One feature removed for prediction |  |  | One feature used for prediction |  |  |
| --- | --- | --- | --- | --- | --- | --- |
|  | Features used | Accuracy (%) | Test statistics | Features used | Accuracy (%) | Test statistics |
| Alpha | 1890 | 65.31<br>(9.34) | $t(9) = -0.81$,<br>$p = .440$,<br>$bf10 = 0.41$ | 60 | 60.22<br>(8.83) | $t(9) = 2.56$,<br>$p = .015$,<br>$bf10 = 5.03$ |
| Beta | 1890 | 66.24<br>(9.09) | $t(9) = 1.22$,<br>$p = .254$,<br>$bf10 = 0.56$ | 60 | 61.78<br>(10.32) | $t(9) = 3.59$,<br>$p = .003$,<br>$bf10 = 19.06$ |
| Theta | 1890 | 66.32<br>(8.87) | $t(9) = 1.35$,<br>$p = .211$,<br>$bf10 = 0.63$ | 60 | 53.58<br>(9.12) | $t(9) = 0.95$,<br>$p = .185$,<br>$bf10 = 0.73$ |
| Connectivity | 180 | 61.55<br>(9.02) | $t(9) = -1.24$,<br>$p = .247$,<br>$bf10 = 0.57$ | 1770 | 66.58<br>(9.34) | $t(9) = 5.42$,<br>$p < .001$,<br>$bf10 > 100$ |
| All together | 1950 | 65.81<br>(9.14) |  |  |  |  |

Decoders were trained either by leaving out one feature set or by using solely one feature set. Performance was averaged across eight minutes and all participants whose decoder performance significantly exceeded chance level ( $n = 10$ ). Performance of reduced decoders was compared to the one encompassing all and those exclusively focusing on one feature with chance level of 50%. Tests were Bonferroni corrected for four tests. Bf10 represents the Bayes Factor.

Additionally, we analyzed the feature weights of the SVMs in the subsample of 10 participants whose decoder performance exceeded chance level. To determine whether feature weights contributed significantly to the decoders’ performance, the feature weights were averaged across cross-validations and bins for these participants and then absolute values were taken. To determine the significance of these averaged feature weights at the group level while correcting for multiple comparisons across the various features, we calculated *p*-values as the proportion of maximum mean feature weights averaged across the aforementioned participants in the feature set from randomized data (*n* = 1000 randomizations) that were equal to or larger than the original feature weights (cf. max-stat approach above). Similarly, feature weights from the ten participants were analyzed at the individual level (Fig. 5). It should be noted that the analysis based on the ten participants with significant decoder accuracies is exploratory in nature and therefore not circular. Consequently, it does not constitute double dipping (Kriegeskorte, Simmons, Bellgowan, & Baker, 2009).

**Figure 5.**
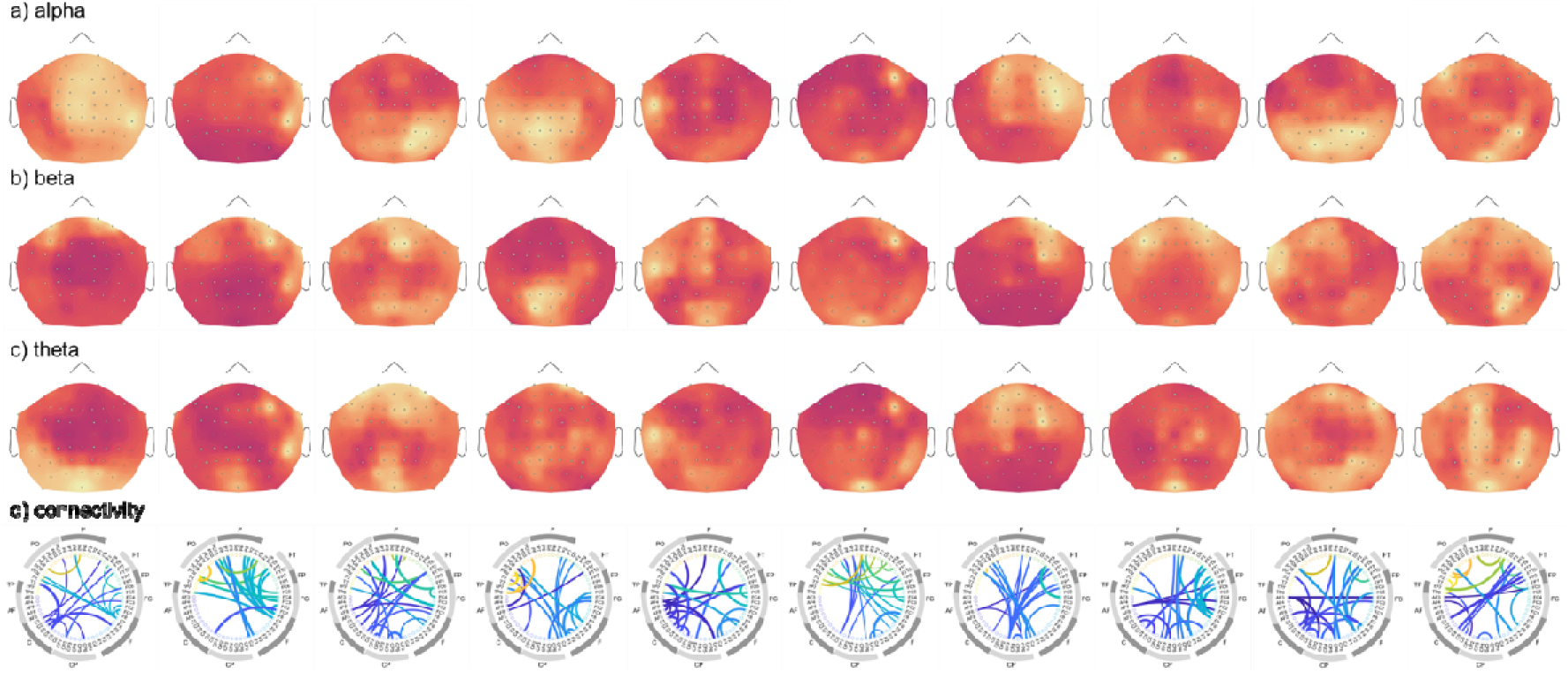
Post-hoc analysis of participant-specific decoder feature weights by channel for participants whose decoder performance significantly exceeded chance level; SVM weights for each feature were extracted. These weights were averaged across bins, cross-validation folds and experimental runs. Absolute weights are presented for each participant for a) alpha, b) beta, c) theta and, d) connectivity. In a), b) and c), brighter colors indicate higher values. Please note that feature weight maps were scaled individually. Each color in the connectivity plot represents one electrode, while the outer circle summarizes electrode subsets. The 20 most informative features from inter-channel connectivity are plotted.

### 2.7 Data availability statement

The datasets and scripts generated and used for analysis in the current study are available in the OSF repository: https://doi.org/10.17605/OSF.IO/QF2HA

## 3 Results

### 3.1 Self-reported rumination (PTQ-S and VAS)

Self-reported rumination increased after the rumination induction and decreased after the distraction induction (Fig. 2a; linear mixed model with significant induction x timepoint interaction for PTQ-S; *F*(1, 69) = 121.77, *p* < .0001, η*_p =_* 0.64). At post-measurements, rumination was significantly higher in the rumination condition (*t*(69) = 10.76, *p* < .0001, *d* = 1.30), whereas at pre-measurements it was significantly lower (*t*(69) = −4.85, *p* < .0001, *d* = −0.58).

Similarly, continuous VAS-reported rumination quickly increased during the rumination induction to a stable plateau and slightly decreased during the distraction induction (Fig. 2B; significant interaction in the linear mixed model between induction and timepoint; *F*(8, 391) = 17.02, *p* < .0001, η*_p_* = 0.26). Thus, VAS-reported rumination differed at each timepoint between both induction conditions (see Fig. 2b). Unexpectedly, but consistent with the PTQ-S pre-measurements, rumination levels already differed slightly, yet significantly, before the induction. This difference resulted from carry-over effects from the previous inductions despite our counter-balanced experimental design: When we analyzed only the first run of each session, we found no difference in rumination between the two induction conditions (see supplemental results, Fig. S1). Consistent with the pre-and post-questionnaire measurements, VAS-reported data indicated that rumination was successfully induced in our sample already from the first minute of rumination induction (i.e., significant difference in self-reported VAS rumination in the rumination induction from each timepoint compared to t0; supplemental results Tab. S2).

### 3.2 Decoding rumination from individual neurophysiological patterns

Having established robust inductions of state rumination, we proceeded to train decoders (i.e., linear SVMs) to distinguish between states of induced rumination and distraction using multivariate EEG patterns, analyzed in one-minute bins across the eight-minute induction periods. The decoders were trained to classify rumination and distraction from training data and then predicted the two mental states from independent test data using cross validation, which allowed to assess the accuracy of decoding.

Decoding accuracy, when averaged **across** all one-minute timepoints, was significantly greater than the chance level of 50% (one-sided one sample *t*-test, *t*(23) = 1.74, *p* =.048, *d* = 0.35 [0.30; 0.40], *bf10* = 1.50), with a mean accuracy of 54.39% (*SD* = 8.76%). Analyzing the accuracies across the bins, we found no significant main effect of timepoint (linear mixed model for accuracies, *F*(8,184) = 1.84, *p* = .073, η*_p_* = 0.01). Post-hoc analyses for each timepoint showed that the decoding accuracies before the induction, as expected, did not differ from chance level (*p* > .05 and Bayes Factor < 1). Descriptively minutes 1, 2, 6 and 7 showed highest decoding accuracies (Fig. 3 and Tab. S4). In line with these findings, accuracies from minutes 1, 6 and 7 differed significantly from minute 0 (supplemental results). However, there was considerable variation in decoding accuracies across participants: Individual accuracies averaged over timepoints ranged from 27.6% to 82.6%. Decoding accuracies differed significantly from chance at the individual level for only 10 participants (Fig. 3; supplemental results Tab. S4). In summary, our decoding results indicate that multivariate EEG patterns contain weak but statistically significant information distinguishing between induced ruminative and distractive states.

### 3.3 Group decoder

The decoders described thus far were trained using individual-level neurophysiological EEG patterns, which may include both generic patterns that are consistent across participants and patterns specific to individuals. To determine whether these decoders primarily exploit person-specific EEG patterns to predict rumination, we trained a ‘group decoder’. This decoder was designed to identify commonalities in the mapping between induced states and EEG patterns across all participants. However, the group decoder achieved a mean decoding accuracy of only 49.59%, which does not differ significantly from the chance level of 50% (*t*(23) = −0.23, *p* = .591, *bf10* = 0.18). Therefore, it appears that the neurophysiological patterns informative for rumination, as used by our individual decoders, are specific to each individual.

### 3.4 Exploratory analysis of the relationship between reported and decoded rumination

Subsequently, we explored the relationship between the neural state of rumination, as quantified using the decoders’ probability estimates of rumination versus distraction, and self-reported rumination throughout the induction phase. At the group level, we observed that the decoders’ probability estimates for rumination diverged from the 50% mark immediately after the onset of the induction, coinciding with an increase in VAS-reported rumination during the rumination induction compared to the distraction induction (Fig. 4a). To analyze this neurobehavioral link more formally, we examined the relationship between the decoders’ probability estimates and the VAS scale ratings over time for each participant. The *z*-transformed linear correlations between the VAS values and the probability estimates differed significantly from zero at the group level (*t*(23) = 2.57 *p* = .009, *bf10* = 6.17; Fig. 4b). Participant-specific correlations between self-reported and decoded rumination are provided in supplementary Tab. S5.

In contrast to the VAS reports, which account for individual time courses of induced rumination, the differences observed pre- and post-induction in the PTQ-S scores – when averaged per induction across sessions and runs – and the mean probability estimates of rumination for each induction did not show a significant correlation (*r* = .10, *p* = .500). Overall, our analyses indicated that there was a weak but significant relationship between decoded and self-reported rumination when taking the participant-specific time courses of induced ruminations into account.

### 3.5 Exploratory analyses of the informativeness of neurophysiological patterns

Our decoding results demonstrated that multivariate EEG feature patterns contain information on ruminative state, and that this information correlates with self-reports. However, it was not clear whether this information was specifically encoded in patterns of the alpha, beta, or theta band, or rather in the connectivity between electrodes, as has been suggested by previous studies on univariate neurophysiological correlates of rumination (Benschop et al., 2021; Ferdek et al., 2016; Keune et al., 2011; Magosso et al., 2021; Putnam & McSweeney, 2008; Umemoto et al., 2021). Therefore, we analyzed the feature weights of decoders from participants whose decoder performance significantly exceeded chance level (*n* = 10 participants with individually significant decoding accuracies). To do so, we chose a ‘virtual lesion’ approach, which evaluates the contribution of each feature set (i.e., alpha, beta, and theta power, as well as connectivity) to overall decoding performance (Kohoutová et al., 2020). This approach trained ‘reduced’ decoders on EEG patterns, which involved omitting one feature set at a time or focusing exclusively on one feature set. We found that the exclusion and inclusion of connectivity between all channels influenced the decoding of rumination: The performance of the ‘reduced’ decoders, which lacked connectivity patterns, was lower than that of a decoder using all patterns, although not significantly (Tab. 1). The decoder trained solely on connectivity showed the best decoding performance, although decoders trained on alpha, beta and theta power also achieved accuracies significantly above chance level.

Additionally, we evaluated the importance of distinct features of the neurophysiological patterns to predict rumination. To do so, we analyzed, at the group level, whether the SVM feature weights among participants with significant decoding accuracies (*n* = 10) differed significantly from chance level using a max-stat (Blair & Karniski, 1993) approach for multiple-comparison correction across features. However, only the beta band activity in channel FP2, F5, F6 and AF8 and the connectivity between channel CP2 and AF3 showed a significant difference from chance, reinforcing the notion that informative features were not consistent across participants. Consistent with this finding, individual-level analysis showed highly variable weight patterns: for example, we found significant results for one participant in the alpha and beta bands in channel P6, while another participant showed significant weights in the beta band, especially in the frontal channels (for detailed results, see supplemental results Tab. S6). Additionally, visual inspection of the feature weight maps (i.e. importance maps) for the alpha, beta and theta bands, as well as the connectivity feature weights, suggested substantial differences across participants (Figure 5).

## 4 Discussion

In this proof-of-principle study, we trained linear decoders to distinguish between states of experimentally induced rumination and distraction based on EEG patterns comprising alpha, beta, and theta power, as well as connectivity between electrodes. The inductions led to significant increases in self-reported rumination as measured in pre versus post assessments (PTQ-S). Continuous self-reported VAS-rumination showed that high rumination levels were quickly achieved within one minute and remained stable across the eight-minute induction period. In contrast, during the distraction induction, VAS-rumination remained at baseline levels throughout the induction period. Across all participants and timepoints, decoding performance significantly differed from chance level (i.e., 50%), albeit at overall rather low decoding accuracies of around 54%. As expected, the EEG patterns did not allow for differentiation between rumination and distraction during resting state before the inductions. Highest decoding accuracies occurred at one, six and seven minutes (all above 55.0%) of the induction period. Post-hoc analyses across all participants and timepoints showed that continuous self-reported VAS-rumination was significantly correlated with decoded rumination. A group decoder trained on patterns across all participants did not exceed chance-level performance. Based on this finding and in line with the notion that rumination patterns are individually different, we found large variability (27.6% to 82.6%) in decoding accuracies between participants, with only 10 out of 24 showing decoding performance above chance level on an individual basis. Additionally, feature weights from the individual significant decoders varied substantially among participants, indicating that these decoders mapped the induced mental states to individually specific neurophysiological patterns. When comparing EEG patterns from different frequency bands and connectivity across participants in more detail, we found that information on the mental states could be mostly attributed to connectivity.

Although our overall decoder performance for state rumination differed significantly from chance level, the accuracies were slightly lower than those observed in studies decoding trait rumination (Aydın & Akın, 2022; Kim et al., 2023). This difference could stem from several factors other than the fundamental distinctions between mental states and traits. For example, our study trained decoders to distinguish between rumination and the complementary neutral state of distraction (Nolen-Hoeksema, 1991), whereas previous research has compared rumination-related EEG features with those of a positive-affect condition (Ferdek et al., 2016), possibly resulting in more pronounced neurophysiological effects. However, despite incorporating a positive-affect condition in pilot tests, we did not observe higher decoding accuracies (supplemental results Tab. S7). The low decoding accuracies for state rumination in our study might also be attributable to the multiple neurocognitive and affective processes involved in rumination, such as executive functions, attention, memory, and abstract semantic reasoning (Watkins & Roberts, 2020; Whitmer & Gotlib, 2013). During rumination, these neurocognitive processes differ from moment to moment within and between individuals, even though the recursive dynamics may form a stable attractor state on a larger timescale. Although these processes may have clear and consistent neural correlates individually, their variable interplay during state rumination likely results in weak and noisy neural signatures of rumination at a global level. Controlling for these processes experimentally is difficult, even with targeted inductions. Additionally, individuals may respond to rumination through cognitive reappraisal, a distinct regulatory strategy from distraction. However, similar to distraction, reappraisal is expected to reduce negative affect by maintaining attention on ruminative thoughts while actively reinterpreting their content (Gross & John, 2003). However, our findings indicate that negative affect remained consistently higher during rumination inductions compared to distraction, whereas positive affect was consistently lower during rumination relative to distraction.

Our findings of interindividual variability in neurophysiological rumination patterns indicate that rumination cannot be conceptualized as a stable, canonical brain state. Instead, the ruminating brain appears to exhibit highly dynamic and individualized activity: Decoding performance significantly exceeded chance level in only 10 out of 24 participants. This relatively small proportion of successful individual decoding outcomes suggests that rumination did not induce strong and reliable neurophysiological signatures within individuals that could be detected through EEG patterns in the alpha, beta, and theta bands and connectivity measures. Additionally, we observed high variability in rumination-related EEG patterns between participants. In the subsample of 10 participants with significant accuracies, we found at group level only very little state rumination patterns in the beta band and the inter-channel connectivity as determined by the decoders’ feature weights; instead, there were highly variable weight patterns across individuals. While previous studies have decoded trait rumination from multivariate patterns across individuals (Aydın & Akın, 2022; Kim et al., 2023) or identified univariate neural features of state rumination that were consistent across individuals (e.g., Ferdek et al., 2016), they did not take account of the neural correlates of rumination within individuals. Our results suggest, however, that the neural basis of state rumination is highly specific to the individual. For example, we observed significant weights in the alpha and theta bands over the temporal cortex in one participant, but increased weights in the beta bands over frontal electrodes in another. Future studies should therefore analyze how individual brain activation correlates with state rumination, speaking to personalized approaches in cognitive and clinical neuroscience (Sui, Jiang, Bustillo, & Calhoun, 2020), as well as computational and precision psychiatry (e.g., Bzdok & Meyer-Lindenberg, 2018; Passos, Ballester, Rabelo-da-Ponte, & Kapczinski, 2021).

Previous studies have rarely examined the dynamics of state rumination. For example, it is unclear over what timescales brain dynamics shift from a non-ruminative to a stable attractor state (Rolls, 2016), a transition that is characteristic of rumination as a recursive process. Given this gap in the evidence, our study was designed to discern how quickly an experimental induction might elicit state rumination, which could occur rapidly (e.g., within a minute) or develop more gradually (e.g., over several minutes). To capture these dynamics, we trained the decoders using one-minute segments of EEG patterns. Within this temporal resolution, our results indicate that the process of rumination begins swiftly, noticeable already within the first minute following the start of induction, but then increases only gradually, eventually reaching a plateau over the induction period of eight minutes. Decoding performance over all participants yielded highest results for minutes 6 and 7, during which time continuously self-reported rumination was also at its peak. Interestingly, our results indicate that self-reported rumination – both in pre-post measurements and continuous monitoring – showed differences right from the start of the induction periods. By comparing only the first induction in each session across participants we were able to demonstrate that these differences arise due to spill-over effects from the previous induction (Fig. S1). This finding indicates that rumination is a stable state over timescales of several minutes.

Based on previous EEG studies investigating neural correlates of trait and state rumination (Benschop et al., 2021; Ferdek et al., 2016; Keune et al., 2011; Magosso et al., 2021; Putnam & McSweeney, 2008; Umemoto et al., 2021), we decided a priori to focus our decoding analysis on spectral EEG patterns in the alpha, beta, and theta bands, as well as on connectivity at the electrode level. Using a ‘virtual lesion’ approach, we found that patterns of connectivity in particular, and to some extent alpha, beta and theta power, provided information on ruminative states. However, our approach did not yield significant decoding results for 14 of the 24 participants, even though this subsample showed strong increases in self-reported rumination (Supplemental Fig. S2). This observation suggests that the few EEG studies that have examined neurophysiological patterns of state rumination (Ferdek et al., 2016; Putnam & McSweeney, 2008) to date may not have fully captured the neurophysiological correlates of state rumination in EEG data. Furthermore, the ability to measure neural signatures of rumination depends on the neuroimaging method. In this study, we used EEG to identify neural signatures of rumination; however, other techniques like fMRI might be better at capturing rumination-related neural activity, such as that within the DMN. For example, fMRI studies (Zhou et al., 2020) have consistently linked the default mode network with rumination, a type of connectivity rarely measured with EEG (although there are exceptions; see e.g., Das, de los Angeles, & Menon, 2022). Despite these limitations, EEG offers advantages in ease of use and affordability compared to fMRI, potentially facilitating the translation of our findings to field studies and clinical settings. The low decoding accuracies seen in a majority of our participants may also result from our choice of a linear SVM decoder. However, our sensitivity analyses indicate that switching to non-linear SVMs did not improve the decoding results (Tab. S8 and Tab. S9).

Our study has at least five limitations: First, we included only moderately to severely depressed participants who exhibited robust changes in self-reported rumination during pre-tests. Thus, our results may not generalize to all individuals with depression, nor to those with other psychiatric disorders in which rumination is an important psychopathological symptom (e.g., Aldao et al., 2010). Second, the accuracy of our decoding of rumination may have been negatively affected by peripheral-physiological signals that were correlated with rumination and thus contaminated the EEG measurements. However, during preprocessing, we cleaned the EEG signals of artifacts related to facial muscles, head movements, and eye movements. Thus, while not absolutely certain, it is highly likely that our decoding results reflect cortical neurophysiological patterns. Third, we continuously assessed subjective rumination using VAS scales. Reporting of rumination every minute may have interfered with cognitive processes, specifically rumination. However, in our proof-of-principle study, we required subjective data to externally validate the decoding results: Only by simultaneously measuring rumination with VAS scales, we could correlate the time course of decoded rumination with subjective rumination. This correlation established an important link between the psychological and neural levels, showing that decoded rumination is psychologically meaningful. Moreover, visually presented items such VAS scales are processed within milliseconds (von der Heydt, 2023) and can quickly be answered. They were applied in the same fashion in the distraction control condition, so that VAS-response-related cognitive or motor processing were controlled. Because we have validated decoded rumination, future studies may use this measure along with or even instead of regular self-reports during the assessment of state rumination, leading to less potential interferences. Fourth, overall decoding accuracies were significant but small. Future studies may increase decoding accuracies. Nevertheless, also small decoding accuracies can help to measure rumination in clinical studies that investigate how interventions affect rumination (Welkerling et al., 2024). Fifth, the large number of features, especially for the connectivity, may have led to variable feature weight maps because individual features are not informed by sufficient data sizes. Therefore, post-hoc analyses regarding the feature weight maps have to be interpreted with caution.

To conclude, our study has shown that neurophysiological spectral and connectivity EEG patterns represent information about individual state rumination. Although the information was weak and encoded in highly variable patterns across individuals, this proof-of-principle study demonstrates that individual dynamic state rumination can be decoded from neurophysiological patterns on a timescale of minutes. These findings may lay the groundwork for future studies investigating the dynamic evolution of neurocognitive and affective processes during state rumination.

### Declaration of Generative AI and AI-assisted technologies in the writing process

During the preparation of this work the authors used ChatGPT (version 4) and DeepL write in order, to improve the readability and clarity of the manuscript. After using this tool/service, the authors reviewed and edited the content as needed and take full responsibility for the content of the publication.

## Supporting information

Supplemental Methods and Results

## Data Availability

The datasets generated and analyzed in the current study are available in the OSF repository,
https://doi.org/10.17605/OSF.IO/QF2HA

## Author contributions

Jana Welkerling: Methodology, Software, Formal Analysis, Investigation, Data Curation, Writing Original Draft, Visualization, Funding acquisition

Patrick Schneeweiss: Resources, Writing Review & Editing

Sebastian Wolf: Conceptualization, Methodology, Funding Acquisition, Writing Review & Editing, Supervision

Tim Rohe: Conceptualization, Methodology, Formal analysis, Funding Aquisition, Writing Review & Editing, Supervision

