## Supplemental Methods and Results for "Decoding ruminative states from neurophysiological patterns"

**Supplemental results and methods**

**Supplemental methods**

*Positive and Negative Affect Schedule (PANAS)*

The German version of the PANAS measures the affect loading on two factors (positive vs. negative affect; PA/NA). 20 adjectives (10 for each factor) were presented to the participants. They were then prompted to provide responses indicating their current feelings on a scale ranging from 1 (“not at all”) to 5 (“extremely”) with respect to the corresponding adjective. Higher values correspond to either more positive or negative affect (Breyer & Bluemke, 2016). Internal consistency for the two dimensions amounts to *α* = .86 (Krohne, Egloff, Kohlmann, & Tausch, 1996).

*Visual Analogue Scales (VAS)*

By adapting the self-assessment manikins from Bradley and Lang (1994), we designed graphical manikins to visualize different levels of rumination and negative versus positive affect. The VAS ranged from 1 to 7. On the VAS for affect, low values corresponded to positive affect and high values to negative. 54 of the 1296 datapoints of self-reported affect based on the VAS scales were missing (4.17%). Missings were handled by averaging the VAS responses per induction and timepoint over the three sessions.

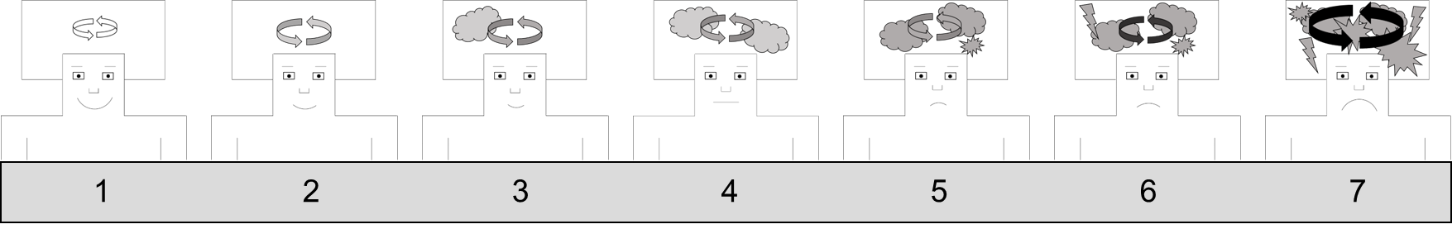

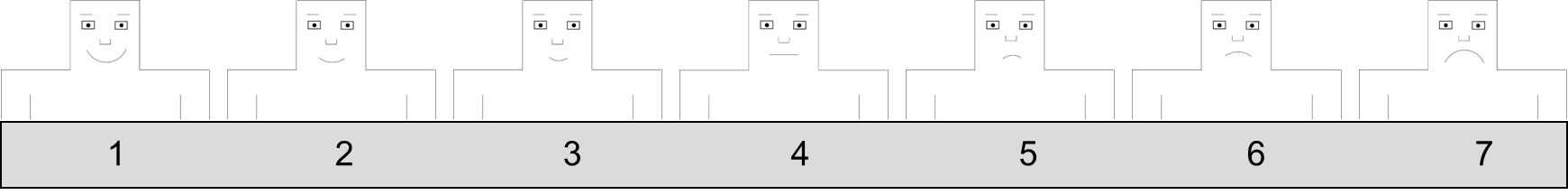

*VAS for rumination and affect.* Higher values in rumination indicate higher rumination, while at the affect scale higher values indicate more negative affect.

**Supplemental results**

*Self-reported affect (PANAS and VAS-scales).*

Self-reported positive affect (PA) decreased after the rumination induction and remained stable after the distraction induction (Fig. S3a; linear mixed model with induction x timepoint interaction for PANAS PA; (*F*(1, 69) = 52.98, *p* < .0001, *η_p_* = 0.43)*_._* At post-induction, PA was significantly higher in the distraction condition (Post: *t*(69) = -6.58, *p* < .0001, *d* = -0.79), while it was significantly lower at pre-induction (*t*(69) = 3.71, *p* < .0005, *d* = -0.45).

Self-reported negative affect (NA) increased after the rumination induction and decreased after the distraction induction (Fig. S3b; linear mixed model with induction x timepoint interaction for PANAS NA; *F*(1, 69) = 103.15, *p* < .0001, *η_p_* = 0.60). At post-induction, NA was significantly higher in the rumination condition (Post: *t*(69) = 9.94, *p* < .0001, *d* = 1.20), while it was significantly lower at pre-induction (Pre: *t*(69) = -4.42, *p* < .0001, *d* = -0.53).

Consistently with the questionnaire self-reports, VAS-reported affect gradually worsened during the rumination induction and remained almost stable during the distraction induction (Fig. S3c; significant interaction in the linear mixed model between induction and timepoint for VAS reported affect; *F*(8, 391) = 13.11, *p* < .0001, *η_p_* = 0.21). Thus, VAS-reported affect differed at each time point between both induction conditions with the exception of minute 0, which was before the inductions (Fig. S3c).

*Decoder Post-hoc Accuracy Analysis.*

Estimated marginal means of the decoding accuracies from minute one (*t*(184) = -2.83, *p* = .005, *d* = -0.21), as well as minute six (*t*(184) = -2.87, *p* = .005, *d* = -0.21) and minute seven (*t*(184) = -2.87, *p* = .005, *d* = -0.21) differed significantly from minute 0.

**Supplemental figures**

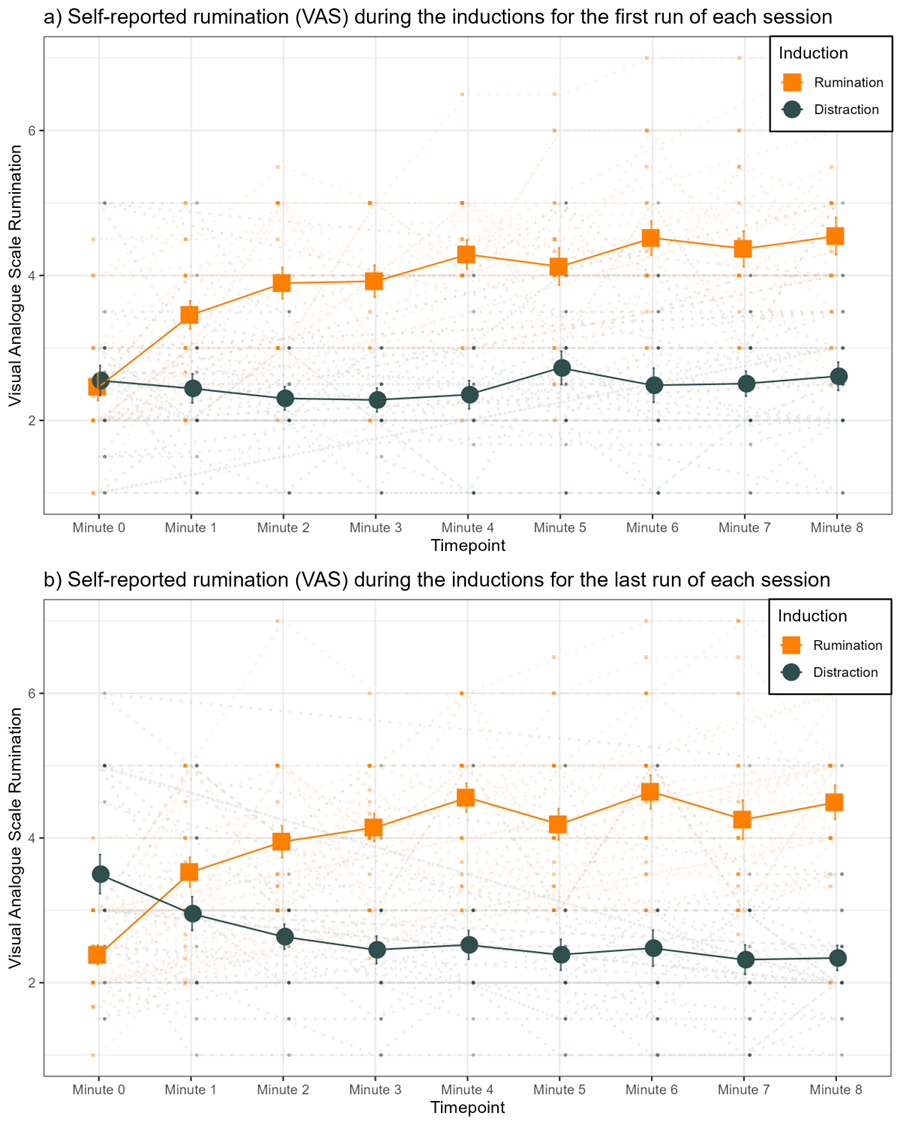

*Figure S1.* *Self-report data from the VAS scales only for the first and last runs in each session for rumination and distraction conditions.* Time course of VAS self-reported rumination (across-participant mean +/- SEM) and individual data in the background for a) only the first run of each session and b) the last run of each session (for the pilot sample last run comprised run two and three). VAS were recorded after the resting state baseline measurement before the inductions (minute 0) and always after 1 minute during the induction (minute 1 – 8).

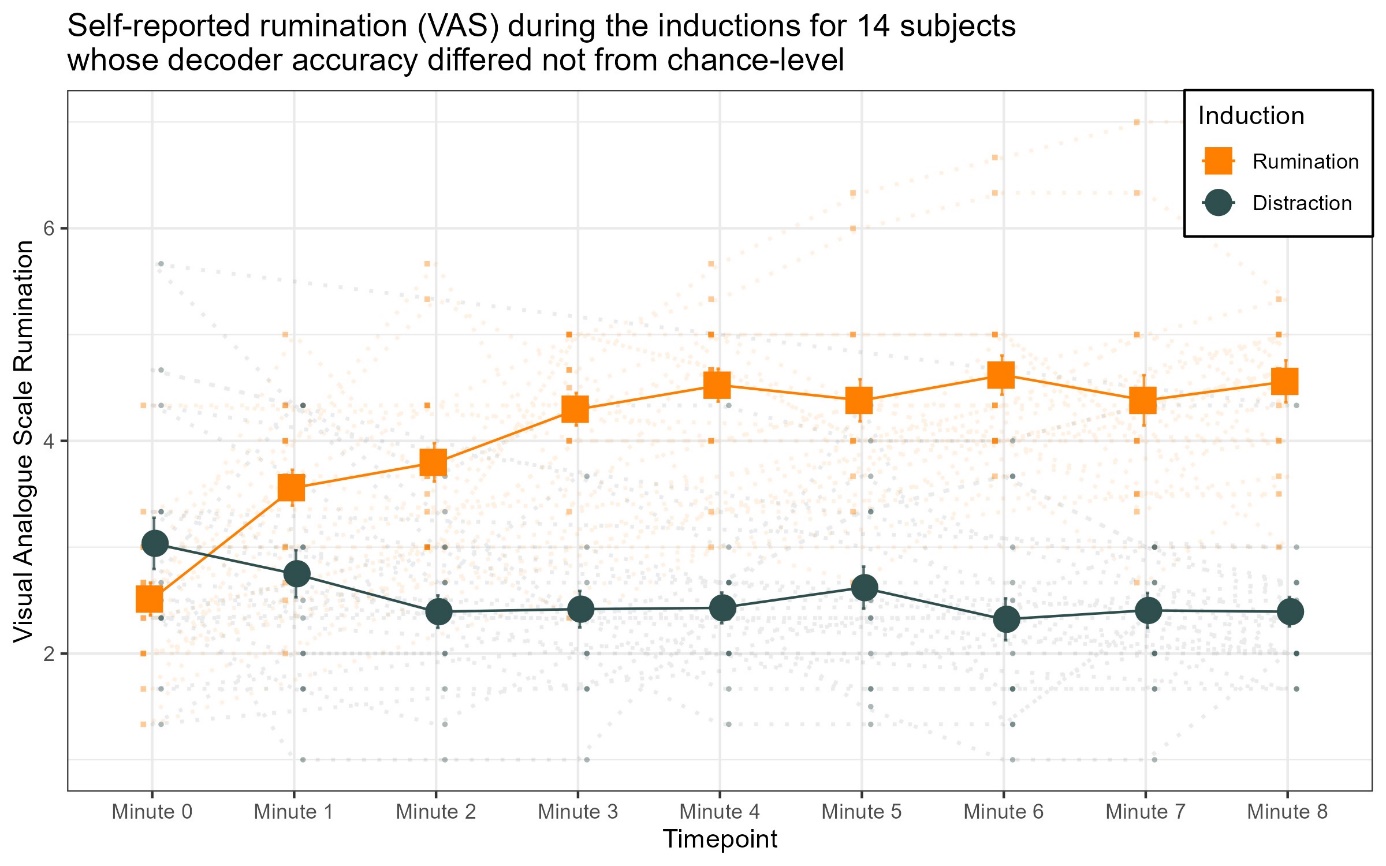

*Figure S2.* *Self-report data from the VAS scales only for the subsample whose decoder performance did not differ from chance-level (n = 14).* Time course of VAS self-reported rumination (across-participant mean +/- SEM and individual data in the background). VAS were recorded after the resting state baseline measurement before the inductions (minute 0) and always after 1 minute during the induction (minute 1 – 8).

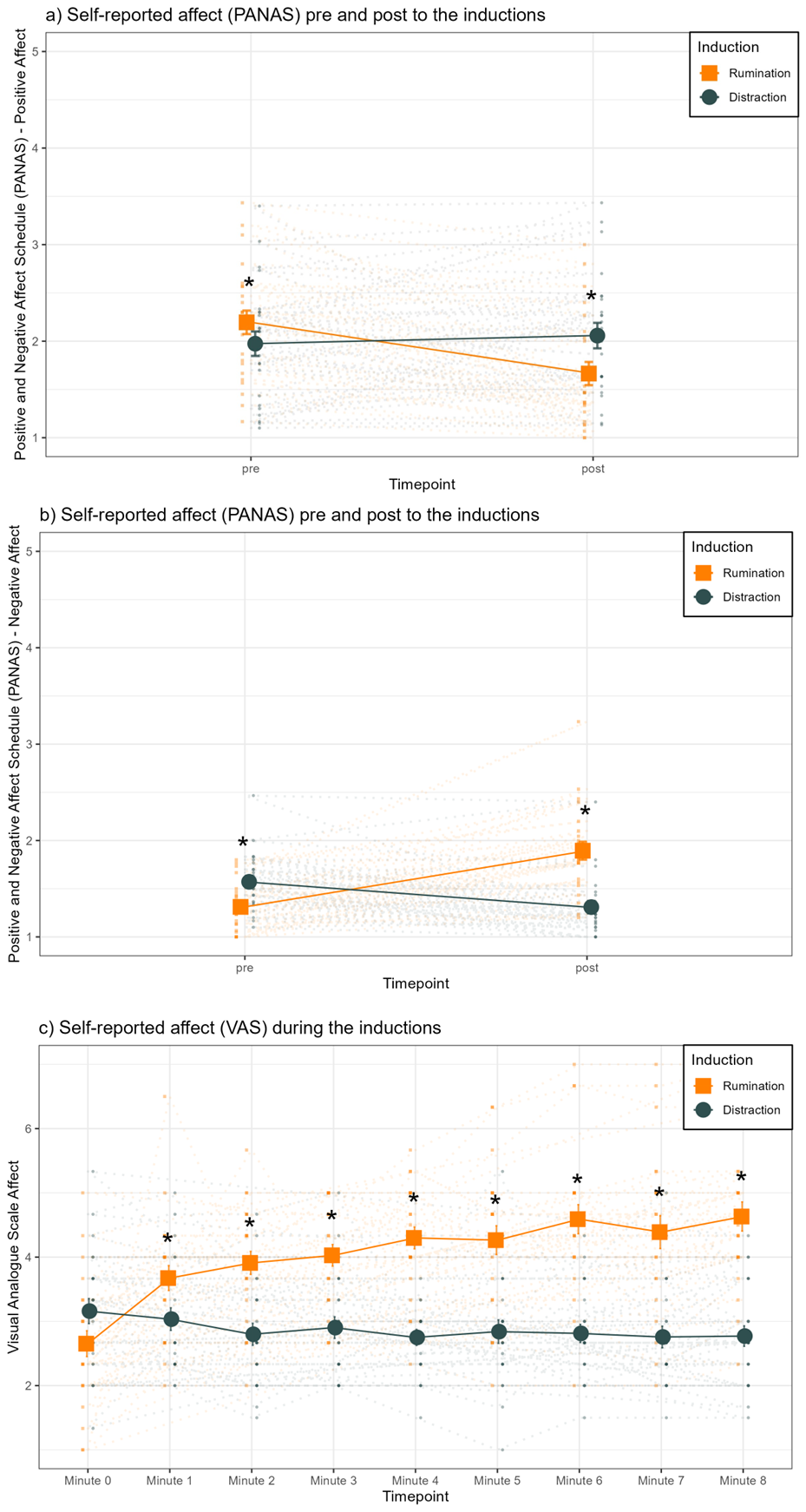

*Figure S3.* *Self-reported affect from the questionnaire and VAS scales for the rumination and distraction conditions.* a) The total score of the PANAS positive affect for pre and post timepoints for each induction (across-participant mean +/- SEM and individual data in the background). b) The total score of the PANAS negative affect for pre and post timepoints for each induction (across-participant mean +/- SEM and individual data in the background). c) Time course of VAS self-reported affect (across-participant mean +/- SEM and individual data in the background). Higher values indicate more negative affect. Asterisks indicate significant results of post-hoc pairwise two-sided *t-*tests between rumination and distraction (see Tab. S3). VAS were recorded after the resting-state baseline measurement before the inductions (minute 0) and always after 1 minute during the induction (minute 1 – 8).

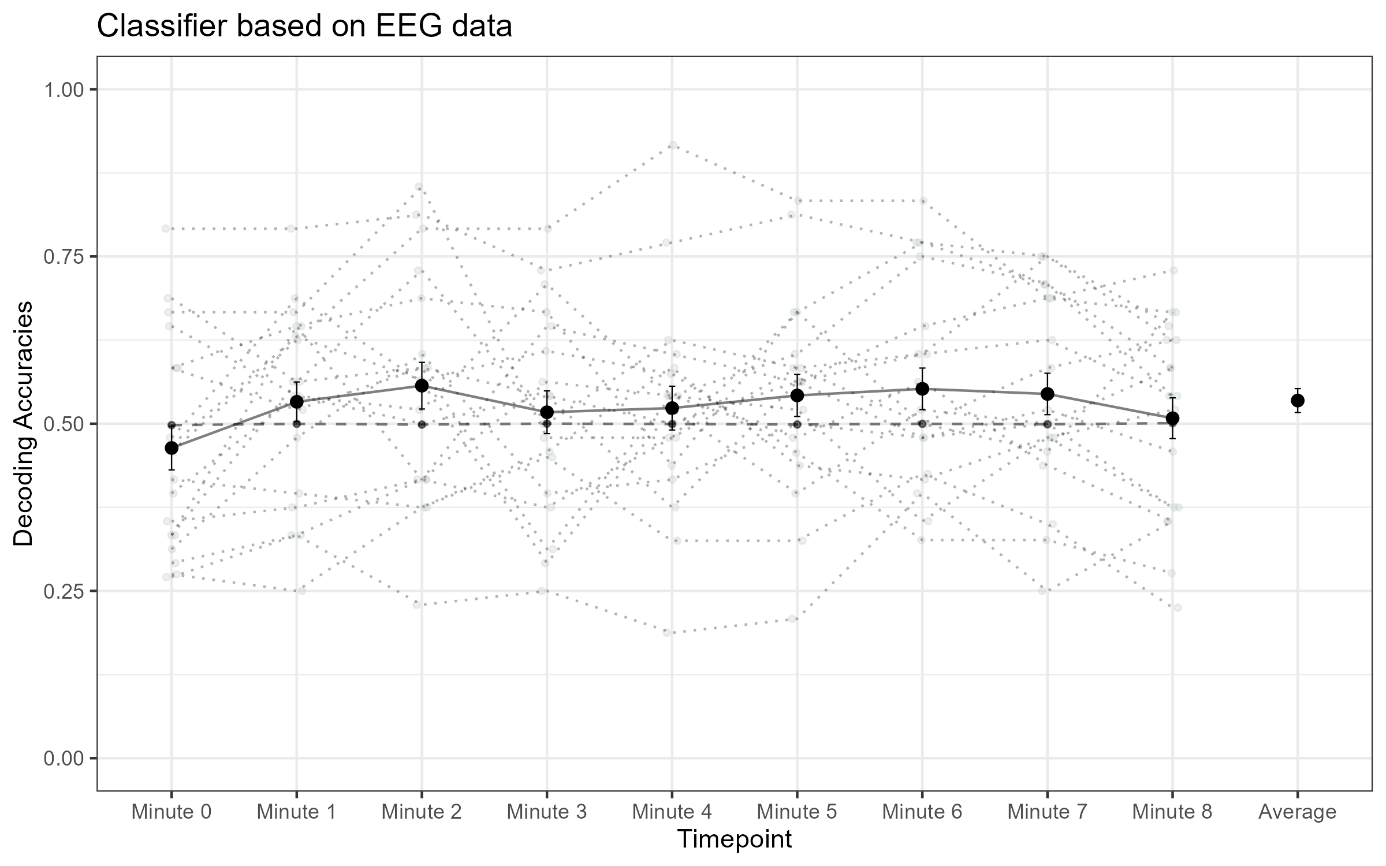

*Figure S4.* *Decoding accuracies of the decoder classifying rumination versus distraction from EEG patterns across the eight-minute induction phase from the main sample (n = 18)*. Decoding accuracies (across-participant mean +/- SEM; individual data dotted in gray) for every one-minute bin and averaged over bins. Chance-level decoding accuracies (black dashed line) were calculated from randomized data (i.e., 1000 random permutations of rumination vs. distraction labels). The results were qualitatively very similar to the complete sample even though the primary analysis (one-sided one-sample *t*-test across bins and participants) was regarding *p*-value and Bayes Factor statistically not significant due to reduced power. However, CI indicate statistic significant results (*t*(17) = 1.19, *p* = .125, *d* = 0.28 [0.23; 0.33], *bf10* = 0.78).

**Supplemental tables**

Table S1. Comparison of average self-reported VAS rumination (across-participant estimated marginal means and SE) for rumination versus distraction inductions at each timepoint.

| Timepoint  (minute) | Rumination | Distraction | Test statistic |
| --- | --- | --- | --- |
| 0 | 2.38 (0.18) | 2.92 (0.18) | *t*(391) = -2.62, *p* < .005, *d* = -0.13 |
| 1 | 3.47 (0.18) | 2.69 (0.18) | *t*(391) = 3.81, *p* < .0005, *d* = 0.19 |
| 2 | 3.80 (0.18) | 2.49 (0.18) | *t*(391) = 6.39, *p* < .0001, *d* = 0.32 |
| 3 | 4.06 (0.18) | 2.35 (0.18) | *t*(391) = 8.36, *p* < .0001, *d* = 0.42 |
| 4 | 4.43 (0.18) | 2.46 (0.18) | *t*(391) = 9.66, *p* < .0001, *d* = 0.49 |
| 5 | 4.18 (0.18) | 2.53 (0.18) | *t*(391) = 8.06, *p* < .0001, *d* = 0.41 |
| 6 | 4.53 (0.18) | 2.44 (0.18) | *t*(391) = 10.23, *p* < .0001, *d* = 0.52 |
| 7 | 4.29 (0.18) | 2.39 (0.18) | *t*(391) = 9.32, *p* < .0001, *d* = 0.47 |
| 8 | 4.45 (0.18) | 2.47 (0.18) | *t*(391) = 9.69, *p* < .0001, *d* = 0.49 |

*At each timepoint, paired two-sided t-tests (Bonferroni corrected for nine tests) were calculated to compare both inductions. Higher values indicate higher self-reported rumination.*

Table S2. Average difference in estimated marginal means of VAS rumination for each timepoint compared to minute 0 for both inductions.

| Timepoint  (minute) | Test statistic rumination induction | Test statistic distraction induction |
| --- | --- | --- |
| 0 vs. 1 | *t*(391) = -5.30, *p* < .0001, *d* = -0.27 | *t*(391) = 1.12, *p* = .263, *d* = 0.06 |
| 0 vs. 2 | *t*(391) = -6.94, *p* < .0001, *d* = -0.35 | *t*(391) = 2.07, *p* = .039, *d* = 0.10 |
| 0 vs. 3 | *t*(391) = -8.23, *p* < .0001, *d* = -0.42 | *t*(391) = 2.75, *p* = .006, *d* = 0.14 |
| 0 vs. 4 | *t*(391) = -10.03, *p* < .0001, *d* = -0.51 | *t*(391) = 2.24, *p* = .025, *d* = 0.11 |
| 0 vs. 5 | *t*(391) = -8.81, *p* < .0001, *d* = -0.45 | *t*(391) = 1.87, *p* = .062, *d* = 0.09 |
| 0 vs. 6 | *t*(391) = -10.51, *p* < .0001, *d* = -0.53 | *t*(391) = 2.35, *p* = .020, *d* = 0.12 |
| 0 vs. 7 | *t*(391) = -9.35, *p* < .0001, *d* = -0.47 | *t*(391) = 2.58, *p* = .010, *d* = 0.13 |
| 0 vs. 8 | *t*(391) = -10.13, *p* < .0001, *d* = -0.51 | *t*(391) = 2.18, *p* = .030, *d* = 0.11 |

*VAS-rumination at each timepoint of each induction is compared with VAS-rumination at minute 0, which belongs to the self-reported VAS rumination before the inductions and after a one-minute resting state measurement, by using paired two-sided t-tests. p-values are Bonferroni corrected for 16 tests. Lower t-values indicate an increase in rumination over time, while higher t-values indicate a decrease in rumination.*

Table S3. Comparison of average self-reported VAS affect (across-participant estimated marginal means and SE) for rumination versus distraction inductions at each timepoint.

| Timepoint  (minute) | Rumination | Distraction | Test statistic |
| --- | --- | --- | --- |
| 0 | 2.65 (0.19) | 3.16 (0.19) | *t*(391) = -2.49, *p* = .013, *d* = -0.13 |
| 1 | 3.67 (0.19) | 3.03 (0.19) | *t*(391) = 3.13, *p* = .002, *d* = 0.16 |
| 2 | 3.91 (0.19) | 2.80 (0.19) | *t*(391) = 5.45, *p* < .0001, *d* = 0.28 |
| 3 | 4.03 (0.19) | 2.90 (0.19) | *t*(391) = 5.51, *p* < .0001, *d* = 0.28 |
| 4 | 4.30 (0.19) | 2.75 (0.19) | *t*(391) = 7.59, *p* < .0001, *d* = 0.38 |
| 5 | 4.26 (0.19) | 2.84 (0.19) | *t*(391) = 6.98, *p* < .0001, *d* = 0.35 |
| 6 | 4.59 (0.19) | 2.81 (0.19) | *t*(391) = 8.71, *p* < .0001, *d* = 0.44 |
| 7 | 4.39 (0.19) | 2.76 (0.19) | *t*(391) = 7.80, *p* < .0001, *d* = 0.39 |
| 8 | 4.63 (0.19) | 2.77 (0.19) | *t*(391) = 9.12, *p* < .0001, *d* = 0.46 |

*At each timepoint, paired two-sided t-tests (Bonferroni corrected for nine tests) were calculated to compare both inductions. Higher VAS-values indicate higher self-reported negative affect.*

Table S4. Individual decoding accuracies for each one-minute bin of the eight-minute induction phase.

|  | Minute 0 | Minute 1 | Minute 2 | Minute 3 | Minute 4 | Minute 5 | Minute 6 | Minute 7 | Minute 8 | Mean |
| --- | --- | --- | --- | --- | --- | --- | --- | --- | --- | --- |
|  | 0.400 | 0.500 | 0.550 | 0.575 | 0.650 | 0.500 | 0.625 | **0.825*** | 0.425 | 0.581* |
|  | 0.646 | **0.917*** | 0.833* | 0.792 | 0.833* | 0.833* | 0.854* | 0.771 | 0.771 | 0.826* |
|  | 0.594 | **0.656** | 0.563 | 0.594 | 0.469 | 0.594 | 0.438 | 0.406 | 0.531 | 0.531 |
|  | 0.500 | 0.450 | 0.375 | 0.450 | **0.625** | 0.475 | 0.550 | 0.550 | 0.475 | 0.494 |
|  | 0.450 | 0.525 | 0.400 | 0.400 | 0.525 | 0.400 | 0.425 | 0.475 | **0.550** | 0.463 |
|  | 0.521 | **0.667** | 0.500 | 0.563 | 0.500 | 0.583 | 0.500 | 0.500 | 0.479 | 0.536 |
|  | 0.792* | 0.792* | 0.813* | 0.729 | 0.771* | **0.813*** | 0.771* | 0.750* | 0.625 | 0.758* |
|  | 0.271 | 0.333 | 0.229 | 0.250 | 0.188 | 0.208 | **0.396** | 0.250 | 0.354 | 0.276 |
|  | 0.354 | 0.375 | 0.417 | **0.563** | 0.542 | 0.479 | 0.563 | 0.438 | 0.354 | 0.466 |
|  | 0.688 | 0.563 | 0.729 | 0.500 | 0.625 | 0.583 | 0.604 | **0.750** | 0.646 | 0.625* |
|  | 0.667 | 0.667 | **0.854*** | 0.479 | 0.479 | 0.667 | 0.771 | 0.708 | 0.542 | 0.646* |
|  | 0.646 | 0.542 | 0.521 | 0.708 | 0.542 | 0.604 | **0.750** | 0.708 | 0.583 | 0.620* |
|  | 0.479 | **0.688** | 0.563 | 0.292 | 0.521 | 0.396 | 0.521 | 0.500 | 0.583 | 0.508 |
|  | 0.468 | **0.630** | 0.565 | 0.609 | 0.574 | 0.457 | 0.326 | 0.326 | 0.277 | 0.471 |
|  | 0.333 | 0.646 | **0.688** | 0.667 | 0.438 | 0.667 | 0.500 | 0.458 | 0.667 | 0.591* |
|  | 0.313 | 0.479 | **0.604** | 0.396 | 0.417 | 0.500 | 0.479 | 0.521 | 0.458 | 0.482 |
|  | 0.458 | 0.625 | 0.792 | 0.792 | **0.917*** | 0.833* | 0.833* | 0.688 | 0.729 | 0.776* |
|  | 0.396 | 0.542 | 0.500 | 0.542 | 0.500 | **0.563** | 0.479 | 0.500 | 0.375 | 0.500 |
|  | 0.417 | 0.396 | 0.375 | 0.458 | **0.583** | 0.438 | 0.417 | 0.479 | 0.521 | 0.458 |
|  | 0.333 | 0.563 | 0.583 | 0.542 | 0.375 | 0.563 | 0.646 | **0.688** | 0.667 | 0.578* |
|  | 0.292 | 0.333 | 0.417 | 0.375 | 0.479 | 0.583 | 0.500 | 0.583 | **0.625** | 0.487 |
|  | 0.583 | **0.646** | 0.417 | 0.646 | 0.604 | 0.563 | 0.604 | 0.625 | 0.542 | 0.581* |
|  | 0.275 | 0.250 | 0.375 | **0.450** | 0.325 | 0.325 | 0.425 | 0.350 | 0.225 | 0.341 |
|  | 0.583 | 0.521 | **0.583** | 0.313 | 0.542 | 0.521 | 0.354 | 0.479 | 0.375 | 0.461 |
| Mean | 0.477 | 0.554° | 0.552° | 0.528 | 0.543 | 0.548 | 0.555° | 0.555° | 0.516 | 0.544* |
| d [CI]  bf10 | -0.14  [-0.08; 0.04];  0.13 | 0.37  [0.00; 0.12];  1.65 | 0.32  [-0.01; 0.12];  1.20 | 0.19  [-0.03; 0.09];  0.52 | 0.28  [-0.02; 0.11];  0.90 | 0.32  [-0.01; 0.11];  1.20 | 0.37  [0.00; 0.12];  1.66 | 0.37  [-0.01; 0.12];  1.65 | 0.10  [-0.04; 0.07];  0.33 | 0.35 [0.30; 0.40];  1.50 |

*Individual decoding accuracies are represented in the cells for every minute. The bins with individually highest decoding accuracies are printed in bold. Additionally, the means across participants per minute are shown in the bottom line of the table. Means per participant across minute (omitting minute 0) can be found in the right column of the table. Decoding accuracies that were significant (p < 0.05) at individual or group level are marked with asterisks. Marginal significant results are marked with circles (p < .10). The overall mean (omitting minute 0) is printed in the right bottom cell.*

Table S5. Individual correlations between probability estimates for rumination predicted by the decoder and self-reported rumination from VAS scales across all timepoints.

| Participant  1  0.418 | Participant 2  0.882 | Participant 3  0.021 | Participant 4  0.055 | Participant 5  -0.028 | Participant 6  0.637 |
| --- | --- | --- | --- | --- | --- |
| Participant 7  0.624 | Participant 8  -0.649 | Participant 9  -0.170 | Participant 10  0.526 | Participant 11  0.600 | Participant  12  0.324 |
| Participant 13  -0.021 | Participant 14  -0.144 | Participant 15  0.359 | Participant 16  0.048 | Participant 17  0.021 | Participant 18  0.040 |
| Participant 19  -0.188 | Participant 20  0.316 | Participant 21  0.279 | Participant 22  0.626 | Participant 23  0.641 | Participant 24  -0.371 |

*VAS values and probability estimates were averaged across sessions and runs for every induction before computing the pairwise correlations for all timepoints of the induction phase per participant between self-reported VAS-rumination and decoded rumination. Pairwise correlations were calculated per subject. Higher coefficients indicate that higher probability estimates are correlated with higher self-reported rumination.*

Table S6. Individually significant SVM feature weights in 10 participants with decoder performances above chance level.

| Participant | Alpha | Beta | Theta | Conn |
| --- | --- | --- | --- | --- |
| 1 |  | FP2, AF7, AF8, F5 |  | PO4/FC1 |
| 2 |  |  |  |  |
| 7 |  |  |  |  |
| 10 |  |  |  |  |
| 11 | T7 |  | T7 |  |
| 12 |  | Oz, F6 | F6 |  |
| 15 |  |  |  |  |
| 17 |  | AF8, F5 |  |  |
| 20 |  | FT7 |  |  |
| 22 | P6 | P6, PO4 |  | F4/FT9, C5/PO8 |

*Weights were extracted per participant, induction, bin and cross-validation. They were then averaged across cross-validation folds and runs. Significance was determined from p-values (p < .05). p-values were computed using a randomization test combined with a max-stat approach to correct for multiple comparison across features. The approach was implemented in each participant.*

Table S7. Individual decoder performance for each one-minute bin of the eight-minute induction phase in a pilot sample for a decoder classifying rumination versus distraction and positive affect.

|  | Minute  0 | Minute  1 | Minute  2 | Minute  3 | Minute  4 | Minute  5 | Minute  6 | Minute  7 | Minute  8 | Mean |
| --- | --- | --- | --- | --- | --- | --- | --- | --- | --- | --- |
|  | 0.453 | 0.391 | 0.422 | 0.344 | 0.547 | 0.422 | 0.453 | 0.594 | 0.453 | 0.453 |
|  | 0.306 | 0.611 | 0.694 | 0.653 | 0.653 | 0.583 | 0.653 | 0.667 | 0.528 | 0.630 |
|  | 0.321 | 0.321 | 0.286 | 0.429 | 0.429 | 0.429 | 0.411 | 0.375 | 0.304 | 0.373 |
|  | 0.219 | 0.297 | 0.266 | 0.266 | 0.219 | 0.141 | 0.328 | 0.406 | 0.250 | 0.271 |
|  | 0.297 | 0.281 | 0.313 | 0.453 | 0.219 | 0.313 | 0.266 | 0.281 | 0.281 | 0.301 |
|  | 0.389 | 0.431 | 0.444 | 0.375 | 0.444 | 0.181 | 0.125 | 0.222 | 0.194 | 0.302 |
| Mean | 0.331 | 0.389 | 0.404 | 0.420 | 0.418 | 0.345 | 0.373 | 0.424 | 0.335 | 0.388 |

*In the pilot sample (n = 6) a decoder was trained based on induced ruminative states in contrast to induced distraction and induced positive affect. Individual decoding accuracies are represented in the cells for every minute. Additionally, the means across participants per minute are shown in the bottom line of the table. Means per participant across minutes (omitting minute 0) can be found in the right column of the table. The overall mean (omitting minute 0) is printed in the right bottom cell. Please note that chance level with 3 label categories is 1/3.*

Table S8. Individual decoding accuracies for each one-minute bin of the eight-minute induction phase from a non-linear polynomial SVM.

|  | Minute  0 | Minute 1 | Minute 2 | Minute 3 | Minute 4 | Minute 5 | Minute 6 | Minute 7 | Minute 8 | Mean |
| --- | --- | --- | --- | --- | --- | --- | --- | --- | --- | --- |
|  | 0.475 | 0.400 | 0.575 | 0.475 | 0.325 | 0.575 | 0.600 | 0.675 | 0.600 | 0.528 |
|  | 0.667 | 0.583 | 0.625 | 0.708 | 0.583 | 0.604 | 0.667 | 0.583 | 0.646 | 0.625 |
|  | 0.656 | 0.531 | 0.500 | 0.500 | 0.500 | 0.438 | 0.438 | 0.500 | 0.500 | 0.488 |
|  | 0.450 | 0.400 | 0.400 | 0.500 | 0.575 | 0.425 | 0.600 | 0.500 | 0.400 | 0.475 |
|  | 0.475 | 0.575 | 0.350 | 0.300 | 0.450 | 0.400 | 0.425 | 0.400 | 0.400 | 0.413 |
|  | 0.500 | 0.667 | 0.667 | 0.604 | 0.646 | 0.500 | 0.500 | 0.438 | 0.292 | 0.539 |
|  | 0.896 | 0.708 | 0.771 | 0.542 | 0.521 | 0.708 | 0.646 | 0.521 | 0.542 | 0.620 |
|  | 0.500 | 0.438 | 0.500 | 0.354 | 0.500 | 0.521 | 0.333 | 0.500 | 0.438 | 0.448 |
|  | 0.458 | 0.583 | 0.375 | 0.500 | 0.500 | 0.479 | 0.604 | 0.500 | 0.500 | 0.505 |
|  | 0.667 | 0.500 | 0.500 | 0.521 | 0.542 | 0.542 | 0.500 | 0.583 | 0.521 | 0.526 |
|  | 0.708 | 0.667 | 0.729 | 0.667 | 0.479 | 0.500 | 0.438 | 0.583 | 0.563 | 0.578 |
|  | 0.604 | 0.688 | 0.521 | 0.563 | 0.521 | 0.688 | 0.542 | 0.500 | 0.563 | 0.573 |
|  | 0.500 | 0.354 | 0.500 | 0.500 | 0.458 | 0.354 | 0.500 | 0.500 | 0.646 | 0.477 |
|  | 0.489 | 0.522 | 0.478 | 0.478 | 0.574 | 0.500 | 0.543 | 0.630 | 0.532 | 0.532 |
|  | 0.500 | 0.458 | 0.625 | 0.646 | 0.667 | 0.667 | 0.479 | 0.479 | 0.500 | 0.565 |
|  | 0.479 | 0.500 | 0.500 | 0.458 | 0.417 | 0.500 | 0.375 | 0.521 | 0.417 | 0.461 |
|  | 0.500 | 0.542 | 0.521 | 0.500 | 0.771 | 0.667 | 0.625 | 0.583 | 0.563 | 0.596 |
|  | 0.500 | 0.479 | 0.521 | 0.542 | 0.521 | 0.500 | 0.521 | 0.521 | 0.333 | 0.492 |
|  | 0.500 | 0.375 | 0.333 | 0.521 | 0.542 | 0.438 | 0.333 | 0.458 | 0.479 | 0.435 |
|  | 0.521 | 0.396 | 0.604 | 0.500 | 0.542 | 0.479 | 0.396 | 0.563 | 0.563 | 0.505 |
|  | 0.500 | 0.542 | 0.479 | 0.500 | 0.500 | 0.500 | 0.646 | 0.542 | 0.354 | 0.508 |
|  | 0.500 | 0.646 | 0.500 | 0.500 | 0.563 | 0.583 | 0.563 | 0.625 | 0.563 | 0.568 |
|  | 0.300 | 0.350 | 0.500 | 0.400 | 0.425 | 0.250 | 0.475 | 0.375 | 0.375 | 0.394 |
|  | 0.500 | 0.500 | 0.625 | 0.438 | 0.375 | 0.458 | 0.396 | 0.500 | 0.458 | 0.469 |
| Mean | 0.535 | 0.517 | 0.529 | 0.509 | 0.521 | 0.511 | 0.506 | 0.524 | 0.489 | 0.513 |

*Individual decoding accuracies are represented in the cells for every minute. Additionally, the averages across participants per bin are shown in the bottom line of the table. Means per participant across minutes (omitting minute 0) can be found in the right column of the table. The overall mean (omitting minute 0) is printed in the right bottom cell.*

Table S9. Individual decoding accuracies for each one-minute bin of the eight-minute induction phase from a non-linear radial SVM.

|  | Minute  0 | Minute 1 | Minute  2 | Minute 3 | Minute  4 | Minute 5 | Minute 6 | Minute 7 | Minute 8 | Mean |
| --- | --- | --- | --- | --- | --- | --- | --- | --- | --- | --- |
|  | 0.550 | 0.500 | 0.525 | 0.475 | 0.550 | 0.525 | 0.625 | 0.650 | 0.475 | 0.541 |
|  | 0.417 | 0.833 | 0.875 | 0.708 | 0.854 | 0.833 | 0.813 | 0.854 | 0.708 | 0.810 |
|  | 0.563 | 0.750 | 0.469 | 0.563 | 0.656 | 0.500 | 0.375 | 0.500 | 0.469 | 0.535 |
|  | 0.575 | 0.425 | 0.600 | 0.575 | 0.625 | 0.575 | 0.700 | 0.550 | 0.600 | 0.581 |
|  | 0.425 | 0.600 | 0.400 | 0.600 | 0.600 | 0.400 | 0.425 | 0.575 | 0.450 | 0.506 |
|  | 0.521 | 0.521 | 0.500 | 0.542 | 0.500 | 0.500 | 0.458 | 0.500 | 0.354 | 0.484 |
|  | 0.813 | 0.708 | 0.792 | 0.833 | 0.771 | 0.792 | 0.708 | 0.708 | 0.792 | 0.763 |
|  | 0.250 | 0.292 | 0.271 | 0.479 | 0.417 | 0.500 | 0.271 | 0.271 | 0.292 | 0.349 |
|  | 0.333 | 0.271 | 0.333 | 0.458 | 0.396 | 0.333 | 0.354 | 0.292 | 0.333 | 0.346 |
|  | 0.729 | 0.667 | 0.708 | 0.542 | 0.458 | 0.604 | 0.625 | 0.563 | 0.521 | 0.586 |
|  | 0.771 | 0.646 | 0.896 | 0.583 | 0.458 | 0.771 | 0.646 | 0.688 | 0.500 | 0.648 |
|  | 0.625 | 0.729 | 0.688 | 0.667 | 0.604 | 0.708 | 0.688 | 0.688 | 0.625 | 0.674 |
|  | 0.417 | 0.708 | 0.542 | 0.333 | 0.458 | 0.438 | 0.521 | 0.479 | 0.667 | 0.518 |
|  | 0.511 | 0.435 | 0.543 | 0.391 | 0.404 | 0.543 | 0.370 | 0.435 | 0.340 | 0.433 |
|  | 0.542 | 0.500 | 0.500 | 0.646 | 0.479 | 0.479 | 0.479 | 0.479 | 0.500 | 0.508 |
|  | 0.438 | 0.646 | 0.500 | 0.396 | 0.583 | 0.646 | 0.563 | 0.542 | 0.500 | 0.547 |
|  | 0.313 | 0.667 | 0.854 | 0.625 | 0.875 | 0.729 | 0.833 | 0.521 | 0.729 | 0.729 |
|  | 0.583 | 0.500 | 0.375 | 0.500 | 0.604 | 0.646 | 0.500 | 0.583 | 0.542 | 0.531 |
|  | 0.625 | 0.521 | 0.625 | 0.563 | 0.583 | 0.625 | 0.750 | 0.583 | 0.688 | 0.617 |
|  | 0.438 | 0.417 | 0.563 | 0.458 | 0.417 | 0.604 | 0.479 | 0.583 | 0.500 | 0.503 |
|  | 0.375 | 0.500 | 0.396 | 0.396 | 0.396 | 0.521 | 0.563 | 0.604 | 0.458 | 0.479 |
|  | 0.667 | 0.521 | 0.438 | 0.604 | 0.583 | 0.583 | 0.625 | 0.729 | 0.667 | 0.594 |
|  | 0.275 | 0.475 | 0.400 | 0.400 | 0.425 | 0.350 | 0.500 | 0.325 | 0.500 | 0.422 |
|  | 0.250 | 0.396 | 0.563 | 0.208 | 0.563 | 0.458 | 0.354 | 0.438 | 0.292 | 0.409 |
| Mean | 0.500 | 0.551 | 0.556 | 0.523 | 0.553 | 0.569 | 0.551 | 0.547 | 0.521 | 0.546 |

*Individual decoding accuracies are represented in the cells for every minute. Additionally, the averages across participants per bin are shown in the bottom line of the table. Means per participant across minutes (omitting minute 0) can be found in the right column of the table. The overall mean (omitting minute 0) is printed in the right bottom cell.*

Table S10. Participants’ sociodemographic information.

|  | N (%) or Mean (SD); [Range] |
| --- | --- |
| Age |  |
| Years | 24.58 (4.63); [18-35] |
| Gender |  |
| Female | 17 (70.8) |
| Male | 7 (29.2) |
| Other | 0 |
| Highest level of education |  |
| None | 0 |
| Secondary | 2 (8.3) |
| Vocational | 0 |
| Highschool diploma | 22 (91.7) |
| Relationship Status |  |
| Married or living with a partner | 11 (45.8) |
| Single, separated, widowd | 13 (54.2) |
| Number of children |  |
| 0 | 24 (100) |
| Current diagnosis (inclusion criteria) |  |
| Moderate depression (F32.1) | 8 (33.3) |
| Recurrent moderate depression (F33.1) | 16 (66.7) |
| Current diagnosis (additional) |  |
| Dysthymia (F34.1) | 5 (20.8) |
| Social anxiety (F40.1) | 1 (4.2) |
| Posttraumatic stress disorder (F43.1) | 2 (8.4) |
| Alcohol abuse (F10.10) | 1 (4.2) |
| Becks Depression Inventory (BDI-II) |  |
| Total Score | 27.46 (5.63); [20-42] |
| Perserverative Thinking Questionnaire (PTQ) |  |
| Total Score | 39.71 (6.82); [30-55] |
| Core features of RNT | 24.00 (5.03); [15-34] |
| Unproductiveness | 8.92 (1.86); [5-12] |
| Mental Capacity | 6.79 (2.47); [4-12] |

*Note: Correlation between symptomatology (BDI) and state rumination (PTQ-S) amounted to r = .22 (p = .16) and between symptomatology and trait rumination (PTQ) to r = .35 (p = .048).*
